# The Impact of Control and Mitigation Strategies during the Second Wave of COVID-19 Infections in Spain and Italy

**DOI:** 10.1101/2021.05.11.21256876

**Authors:** Marco De Nadai, Kristof Roomp, Bruno Lepri, Nuria Oliver

## Abstract

European countries struggled to fight against the second and the third waves of the COVID-19 pandemic, as the Test-Trace-Isolate (TTI) strategy widely adopted over the summer and early fall failed to effectively contain the spread of the disease. In this paper, we shed light on the effectiveness of such a strategy in two European countries (Spain and Italy) by analysing data from June to December 2020, collected via a large-scale online citizen survey with 95,251 answers in Spain and 43,393 answers in Italy. Through our analysis, we identify several weaknesses in each of the three pillars of the TTI strategy: testing, tracing and isolating. Moreover, we analyse the respondents’ self-reported behaviour before and after the mitigation strategies were deployed during the second wave of infections. We find that the changes in the participants’ behaviour were more pronounced in Italy than in Spain, whereas in both countries, respondents reported being very compliant with individual protection measures, such as wearing facial masks or frequently disinfecting their hands. Finally, we analyse the participants’ perceptions about their government’s measures and the safety of everyday activities and places regarding the risk of getting an infection. We find that the perceived risk is often gender- and age-dependent and not aligned with the risk level identified by the literature. This finding emphasises the importance of deploying public-health communication campaigns to debunk misconceptions about SARS-CoV-2. Overall, our work shows the value of online citizen surveys to quickly and cheaply collect large-scale data to support and evaluate policy decisions to contrast the spread of the disease.

## INTRODUCTION

Since March of 2020, Europe has been fighting the coronavirus pandemic. The first wave of infections during March and April of 2020 led to a significant excess in mortality rates [1–4] and caused unprecedented pressure in the healthcare systems of many European countries. After applying severe confinement measures from mid-March until June 2020, the first wave of SARS-CoV-2 infections was controlled in Europe [5–7]. Companies and retail activities reopened, and the majority of non-pharmaceutical interventions were lifted [8, 9]. To prevent a potential second wave of infections, most of the European governments focused on a control strategy called Test-Trace-Isolate (TTI) [10–13], aimed at detecting and isolating all infected individuals as early as possible to limit their ability to infect other people and thus reduce the number of COVID-19 cases. Given the emergence of second and third waves of COVID-19 infections in Europe and the necessary implementation of pandemic mitigation interventions, questions were raised about the efficacy of the implemented TTI control strategies. Unfortunately, governments do not typically collect (or are unable to share) detailed statistics about each of the three pillars –namely, Trace-Test-Isolate– of this strategy, making it challenging to identify the strengths and weaknesses of the implemented measures to control the spread of coronavirus.

This manuscript investigates the effectiveness of the control strategies and mitigation interventions implemented in Spain and Italy between June and December of 2020 by analysing data collected via a large-scale online citizen survey with 95,251 answers in Spain and 43,393 answers in Italy. Spain and Italy are two of the most affected European countries with more than 65,000 reported COVID-19 cases and over 1,660 COVID-19 related deaths per million people to date [14]. We divide our analysis into two phases: a first phase between the end of the first wave and the start of the second wave of coronavirus infections (between June and September 2020) and a second phase, corresponding to the second wave of SARS-CoV-2 infections (between October and December 2020).

Through our analysis, we identify several weaknesses in each of the three pillars of the TTI control strategy. Moreover, we shed light on the participants’ behaviour before and after the mitigation strategies were deployed during the second wave of infections. We find that respondents reported being compliant with individual protection measures (e.g. facial mask wearing, hand disinfecting, keeping physical distance) in both countries, and the reported changes of behaviour were larger in Italy than in Spain. When analysing the participants’ perceptions about their governments’ measures and the risk of getting a coronavirus infection associated with common places and activities, we find that perceived risk is in some cases gender-and age-dependent and not aligned with the risk level identified by the literature. Our work illustrates the value of citizen surveys as a cheap and efficient tool to quickly collect large-scale population data about people’s perception, situation and self-reported behaviour during a pandemic.

## RESULTS

We analyse 138,644 answers to an anonymous online survey called COVID19ImpactSurvey [15], which two of the authors designed and launched on March 28th 2020. The survey consists of 26 questions that ask participants about their demographic and household information, their social behaviour and adopted protection measures to prevent a coronavirus infection (e.g. face mask usage), their ability to isolate, their willingness to get tested and vaccinated, and their perception about the adopted government measures and the risk of infection associated with different activities/places (e.g. restaurants). Our results are based on analysing a subset of the 26 questions as per Table S1 in Supplementary Information (SI). We focus on the answers collected between the end of June and the end of December 2020 in Spain and Italy, namely 95,251 and 43,393 answers, respectively. Each day, the survey collected, on average, 491 anonymous answers in Spain and 217 ones in Italy. Users were required to be at least 18 years old.

The gender and age of each participant are not proportional to those of the general population of Spain and Italy. Thus, we follow Oliver *et al*. [15] and reweight the answers to match the official demographic distribution as those of Spain, reported by the Spanish National Institute of Statistics (INE) and the Italian National Institute of Statistics (ISTAT) in 2020. We also filtered entries with inconsistent answers (4% and 6% of answers in Spain and Italy, respectively) and entries that appeared to have been answered too fast (2% and 4% of answers in Spain and Italy, respectively) or too slow (2% and 3% of answers in Spain and Italy, respectively). In total, 8% and 13% of the answers were discarded in Spain and Italy, respectively. We refer to the Material and Methods section for additional details. All answers are categorical or binary, thus we report the percentage of participants who selected each response and compute the 95% confidence intervals through the margin of error.

We temporally divide our analyses according to two phases: (1) *Phase I -new normality* and (2) *Phase II -second wave. Phase I -new normality* corresponds to the time period between June and October of 2020 (Italy: July 31st -October 26th, Spain: June 21st -October 25th). During this phase, both Spain and Italy lifted most of the non-pharmaceutical interventions deployed since March of 2020 and only imposed local measures. Hence, this period is referred to as the *new normality*. Most of the activities were re-established as they were before the start of the pandemic except for requiring facial mask wearing in indoor/outdoor public spaces, defining limitations on large gatherings and light-weight restrictions such on the occupancy of gyms, restaurants, theatres and cinemas. *Phase II -second wave* corresponds to the period between October and December 2020 (Italy: October 26th -December 31st, Spain: October 25th -December 31st), when the second wave of the coronavirus pandemic took place in both countries. During this phase, the countries implemented a range of non-pharmaceutical interventions to contain the spread of the disease, including mobility restrictions, partial closings of restaurants, bars, coffee shops and gyms, and cancellations or limits on the size of large events (e.g. sports events, weddings, funerals) and on the number of people in small family/social gatherings. We refer to the Methodology Section for additional details.

We first report our findings through the lens of the survey related to the Trace-Test-Isolate control strategies implemented in Spain and Italy during the *new normality* phase, followed by an analysis of the reported changes in the respondents’ behaviour as a consequence of the implemented mitigation interventions deployed during the *second wave* phase.

### A. Effectiveness of the Trace, Test and Isolate Control Strategies in Spain and Italy

#### 1. Trace

Contact tracing is typically performed by case investigators and contact tracers, specifically trained to locate and talk with people who have tested positive for coronavirus. The focus is on understanding how people got infected, identifying the close contacts they had up to that point and determining which of those should be contacted to recommend testing and isolation if necessary.

To estimate the level of contact tracing implemented in each country, we analyse the responses of participants who report having had a close contact with someone infected by COVID-19 (Q11 in SI Table S1). Following the World Health Organization’s definition, a close contact is someone who has been within 1.5 meters of an infected individual for at least 15 cumulative minutes over a 24-hour period [16]. From those, we determine the percentage who also report having been called by a contact tracer (Q11_1 in SI Table S1). Although Q11 was not originally designed to measure trace effectiveness and might underestimate the number of traced contacts, it provides a valuable proxy of the number of close contacts and tracing efficacy and enable a comparison of the values across time and in the two countries.

Regarding the percentage of those who reported having had a close contact with a positive individual and were contact traced, we obtain similar results in Spain and Italy. We find that, on average, only 29% of respondents reported having been called by the contact tracers (Italy: 25%, Spain: 33%). Figure 1A shows the results of the two countries aggregated by the number of weekly contacts (Q9 in SI Table S1). Interestingly, we observe that the two countries were especially weak in tracing participants with the highest number of close contacts (50+). We also investigate contact tracing among those who reported having tested positive for coronavirus when filling out the survey (Q24 answers a-b-d-f and Q24_3 in SI Table S1). We find that 75% of participants reported having been contacted by contact tracers (Italy: 78%, Spain: 74%); 62% declared that some of their close contacts were traced (Italy: 62%, Spain: 62%), and 42% reported that their close contacts were also tested for coronavirus (Italy: 33%, Spain: 46%) (see SI Figure S5).

**FIG. 1.**
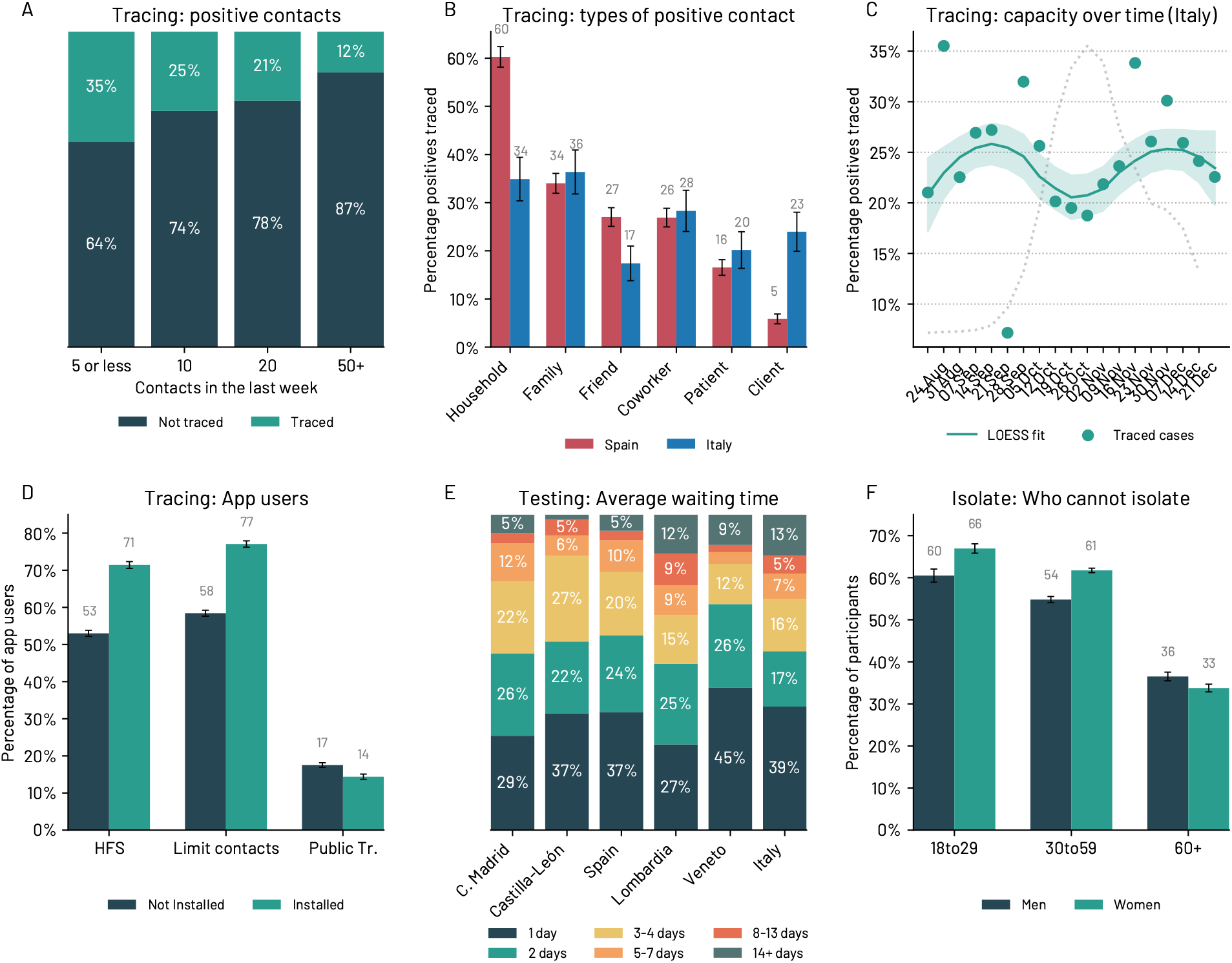
The Tracing-Testing-Isolate (TTI) control strategy during the Phase I - *new normality* in Italy and Spain. A) The majority of respondents who had contact with a positive case report were not contacted by the health authority. B) Percentage of contacts traced for different categories of contacts. Spain focused the tracing activity within the household, where more than 60% of the people reported having been contact traced. C) In Italy, after summer, the tracing capacity seems to be decreased as the number of positive contacts increased. D) Respondents who installed the contact tracing apps are, on average, more prone to comply with the washing Hands, Face masks, making Space (HFS) recommendations^a^). Moreover, app users are more prone to limit their close contacts. E) Average waiting time to get the test results in Spain, Italy and two exemplar regions in the two countries. F) Roughly 50% of the respondents declared having problems self-isolating if needed. The barriers are particularly evident among the youth and women. In B), C), D) and F) we report the 95% confidence interval.

We observe significant differences between Spain and Italy when we consider the *type of contact* (Q9_1 in SI Table S1) traced. According to our data, coworkers and friends are the most common types of close contacts reported by infected individuals (see SI Figure S3). However, Figure 1B shows that less than 25% of these contacts were traced, while the contact types that were most frequently traced are within the household and family. Notably, in 60% of the cases, the household contacts were contact traced in Spain, while the other types of the close contacts were traced less than 40% of the time.

We also computed the trend of the percentage of participants who reported both having had a close contact with a positive case and having been traced by the health authority in Italy by means of a symmetric Locally Estimated Scatterplot Smoothing (LOESS) model [17]. Figure 1C depicts such trend together with the daily number of coronavirus infections. The Figure illustrates that, in Italy, the percentage of traced contacts per infected individual decreased as the number of positive COVID-19 cases increased, suggesting a saturation of the contact tracing capacity.

To complement manual contact tracing efforts, many governments in the world launched smartphone apps to automatically trace and inform people who had been in close contact with an infected individual, following the theoretical model proposed by Ferretti *et al*. [18]. Spain and Italy launched in the summer of 2020 their Bluetooth-based digital contact tracing apps based on the GAEN (Google and Apple Exposure Notification) interface [19]: RadarCOVID launched on August 14th 2020 in Spain, and Immuni launched on June 15th 2020 in Italy. While the apps had reached 6.3 and 10.1 million users respectively by the end of December of 2020, their penetration was not evenly distributed demographically. According to our data, in Italy, more than 50% of male respondents aged 18-29 years old, 41% of male respondents aged 60+ years and 47% of female respondents aged 60+ reported having the app installed (Q20 in SI Table S1). These figures are higher than the officially reported adoption statistics for Italy, reflecting a technology bias in our sample. In Spain, we identify a gender and age difference: on average, women do not install the app as much as men do, and younger participants are less likely to have the app installed than older individuals. We refer to SI Figure S6 for additional details.

Interestingly, we observe that app users adopt, on average, significantly more countermeasures to prevent the spread of coronavirus than non-app users (Q20 in SI Table S1). App users are more likely than non-app users (71% vs 53%) to comply with the Hands/Face/Space (HFS) recommended measures –i.e. wash/disinfect their hands, wear face masks and maintain physical distance with other people; they tend to limit their close contacts more (77% vs 58%) and use less frequently public transportation to commute (14% vs 17%). We refer to SI Figure S13 for additional details.

Our data also allows us to understand the added value of the apps to trace contacts with positive individuals (Q11 and Q11_1 in SI Table S1). We observe that only 2% of respondents who reported having had a close contact with an infected individual received a notification from the app (Italy: 3%, Spain: 1%). Moreover, only 1% of respondents (Italy: 2%, Spain: less than 1%) had close contact with a positive case discovered through the app. Of those, only one case in Italy got tested, while none of them got tested in Spain out of the 698 people who got a notification, according to the survey (Q11_1 in SI Table S1).

#### 2. Test

The second TTI component entails promptly testing all suspected individuals with a coronavirus infection due to having had close contact with an infected case and/or exhibiting symptoms attributable to COVID-19.

On average, in Spain and Italy, 62% and 57%, respectively, of respondents reported receiving the test results within 48 hours from taking the test (Q24 answers a-b-d-f and Q24_3 in SI Table S1). However, we observe significant differences per administrative region. In the Autonomous Community of Madrid (Spain) around 54% of tests took 48 hours and 9% of tests took eight days or more, while in Castilla-Leon (Spain) 59% of tests were reported within 48 hours and only 7% took eight days or more. Similarly, in Lombardia (Italy) 53% of tests took 48 hours to be reported and 22% of the tests took eight days or more. Veneto (Italy) was more efficient than Lombardia: only 12% of the tests required eight days or more to be processed.

In addition, we have evidence that the time required to get a test result decreased over time in Spain and Italy. In Italy, the average waiting time to receive the test results in October was 4.0 days, while in December it went down to around 3.5 days. Similarly, in Spain, it changed from 3.1 days in October to around 2.8 days by the end of December (see SI Figure S9). This decrease in waiting time is also confirmed in Italy by the official data shared by the Italian health authority [20]. Note that the shared data might be noisy and influenced by how it is reported to the health authority [21]. Thus, it is not possible to compare the absolute values between our survey and the official data. However, we observe that over time, Italian delays in reporting COVID-19 test results seem to decrease from 7.5 days on average in September 2020 to around three days at the end of December 2020. As expected, we see a weekly trend where there are significantly fewer tests on the weekends and holidays. We refer to SI Figure S10 for additional details.

#### 3. Isolate

Typically, European governments ask patients with COVID-19 compatible symptoms to self-isolate at home [22] in addition to asking all positive cases and those who had had close contact with an infected individual until obtaining their COVID-19 test results. However, we find that a non-negligible percentage of respondents report being unable to self-isolate due to various factors (Q19 in SI Table S1). Figure 1F shows that more than 54% of participants aged 18-59 years old would be unable to isolate effectively. Fortunately, older (60+ years old) and more vulnerable participants report having fewer self-isolation problems (less than 38% report being unable to self isolate). Our data also shows that more than 7% of respondents would have financial difficulties (Italy: 9%, Spain: 7%), are afraid of stigmatisation (Italy: 11%, Spain: 7%), and/or find isolation psychologically impossible (Italy: 10%, Spain: 8%). Moreover, we identify significant age and gender differences in the reported barriers. Women aged 30 to 59 years old are the most likely demographic group to report an inability to self-isolate due to having to take care of children (29% in Italy and 28% in Spain). Interestingly, men of the same age group are significantly less likely to report facing the same barrier (14% in Italy and 13% in Spain). Similar differences are present between women aged 18 to 29 (10% in Italy and 13% in Spain) and men aged 18 to 29 (4% in Italy and Spain) years old.

The youth (aged 18-20 years old, and particularly women) is the most likely demographic group to report psychological barriers to self-isolation: 22% of youth in Italy and 19% of youth in Spain report being afraid of stigmatisation due to coronavirus; and 28% of youth in Italy and 21% of youth in Spain report not being able to self-isolate due to psychological factors. More than 40% of Italian women aged 18 to 20 years old declare that it would be psychologically impossible for them to be quarantined for at least ten days, while more than 24% are afraid of stigmatisation. Similar results apply to young respondents in Spain.

Financial and labour barriers that prevent self-isolation are the most frequent among those aged 21 to 39, with significant differences compared to other age groups. Conversely, the elderly (aged 60+) is the most likely demographic group to report being able to self-isolate, with significant differences compared to other age groups. For example, in Italy, 18% of the elderly vs 35% for those aged 18-59 years old report not being able to self isolate. Similar results are obtained for Spain. We refer to the SI Figure S7 for additional details.

### B. Community engagement

The cooperation of the population by complying with social distancing and personal protection measures is essential to control the transmission of coronavirus and maintain a low reproduction number [23]. Thus, we analyse the participants’ perception of the risk of getting a coronavirus infection associated with performing different activities and/or going to various places (Q15 in SI Table S1). We also analyse the personal protection measures they usually adopt to prevent a SARS-CoV-2 infection (Q20 in SI Table S1) and whether they telework regularly (Q17, Q17_1 and Q18 in SI Table S1).

We find significant differences in the behaviour of participants in the two countries. Hence, we report the results for each country separately. Figure 2A suggests that Spanish respondents think that fewer places are safe than Italians think. For example, only 31% of Spaniards consider restaurants to be places with a low risk of getting COVID-19 compared to 50% of Italian respondents. Similarly, schools are perceived as significantly safer by Italian (39%) than by Spanish (27%) participants. Moreover, we identify significant gender-based differences in the perception of risk. For example, men are more likely than women to consider that individual sports (Italy: 74% vs 65%, Spain: 79% vs 73%), restaurants (Italy: 54% vs 47%, Spain: 35% vs 29%) and having friends at home (Italy: 40% vs 33%, Spain: 27% vs 22%) can be performed with low risk of getting a coronavirus infection. This perception of safety also changes with time: as the cumulative incidence of COVID-19 cases rises, we observe a consistent increase in the perception of risk of all activities and places in both countries. However, the ranking of the activities/places remains the same throughout the study: air travel is consistently considered the activity with the highest risk of getting infected with COVID-19 and practising individual sports the activity with the lowest risk of infection. We refer to the SI Figure S11 and Figure S12 for additional details.

**FIG. 2.**
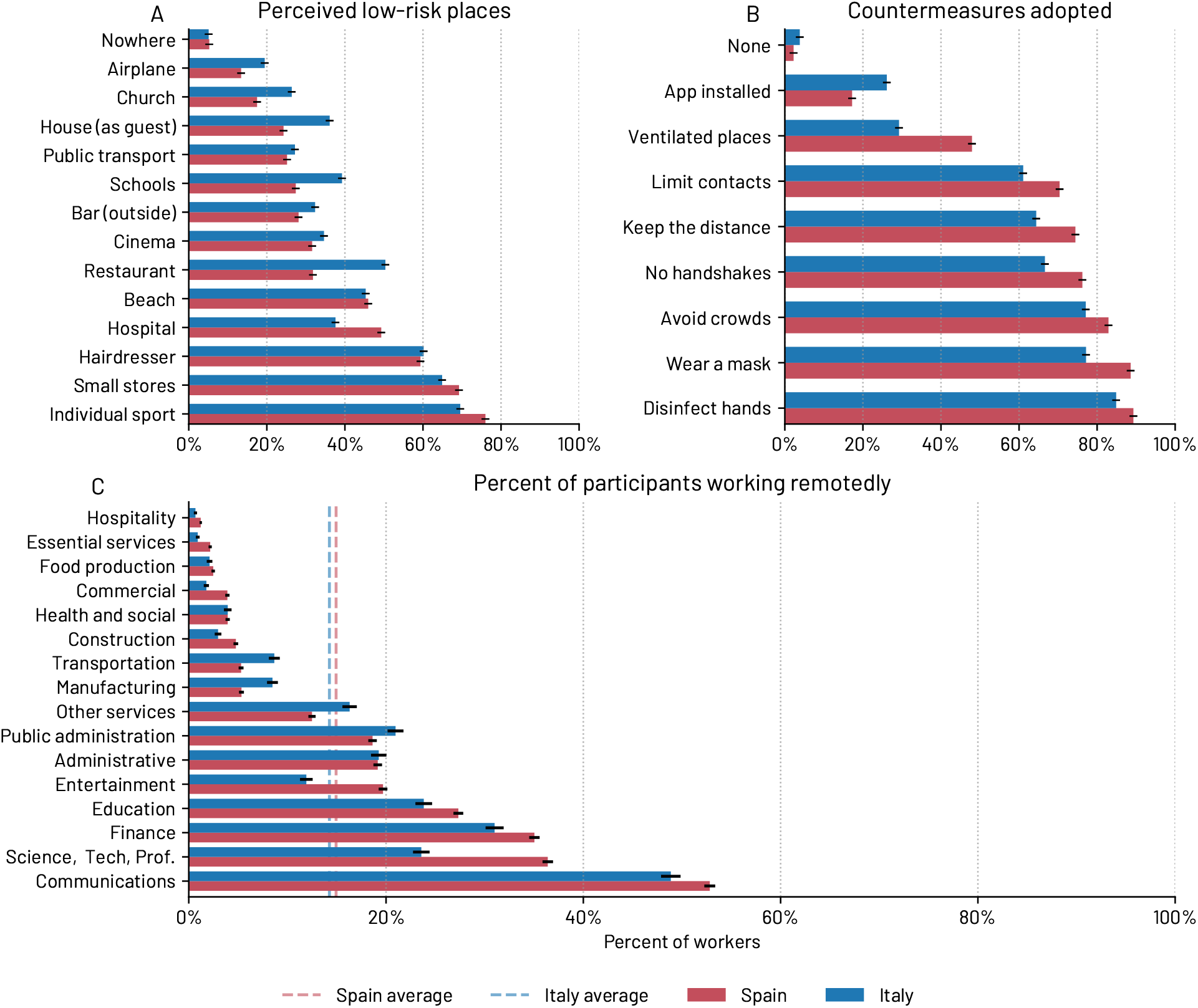
Human behaviour in the Phase I - *new normality*. A) Perception of safety (in terms of risk of getting a COVID-19 infection) for different activities/places in Spain and Italy. B) Individual COVID-19 protection measures adopted in Spain and Italy. C) Percentage of workers that report teleworking per sector, in Spain and Italy. In this figure, we report the 95% confidence intervals.

Figure 2B shows the personal protection measures adopted to prevent a coronavirus infection. Again, we observe that Spanish respondents are more cautious about COVID-19 and adopt more countermeasures than Italians in all areas but the app installation, where there are significantly more Italian respondents with the app installed than Spanish respondents (Italy: 26%, Spain: 17%). The most notable difference is found in the adoption of ventilation: 48% of Spanish vs 29% of Italian participants report ensuring good ventilation indoors.

We identify significant gender and age differences, with women and older respondents being generally more compliant and cautious than men and the youth. For example, women are more likely than men to report avoid shaking hands (Italy: 72% vs 61%, Spain: 78% vs 74%), limit their contacts (Italy: 66% vs 56%, Spain: 75% vs 66%) and disinfect hands (Italy: 89% vs 80%, Spain: 92% vs 86%). We report more information in SI Figure S13 and Figure S14.

Finally, most countries deployed public communication campaigns and programs to promote telework and hence reduce the risk of infection at work. Figure 2C shows the percentage of participants who report remote working in the new normality phase in Spain and Italy. The categories that have more remote workers are Communications, Science/Tech/Professional and Finance, which are also amongst the most digital sectors [24]. Although the average is similar in both countries (around 15%) and significantly larger than the values before the COVID-19 pandemic (*∼*5% in Italy and *∼*6% in Spain in 2019 according to a recent European report [25]), we observe significant differences per sector per country, particularly in the Communications (52% vs 48%), Science/Tech/Professional (38% vs 23%) and Entertainment (19% vs 11%) sectors. In these sectors, a significantly larger percentage of Spanish respondents report teleworking when compared to Italian respondents, which might be a consequence of Spain having a larger Digital Technology Integration Index when compared to Italy (42 vs 33 according to [25]).

## MITIGATION INTERVENTIONS: BEHAVIOUR AND PERCEPTION

### C. Human Behaviour and Mitigation Interventions

In this Section, we compare the participants’ self-reported behaviour before and after the mitigation non-pharmaceutical interventions were deployed to combat the second wave of coronavirus infections in each country.

With the rise of confirmed COVID-19 cases in Italy in early October, the Italian Prime Minister imposed on October 7th 2020, the compulsory use of face masks in public spaces [26]. Between the 13th and the 26th of October, stricter rules were implemented, such as in the opening hours and maximum allowed capacity of restaurants and bars [27–29]. Museums, theatres, cinemas, gyms and swimming pools were closed. After one month of rise of confirmed COVID-19 cases, the government defined a three-tiered system on November 6th 2020 [30]. Each region was assigned a tier depending on several quantitative indicators: i) the level of transmission (e.g. the effective reproduction number, the number of positive cases in the past two weeks, the number of novel outbreaks), ii) the resilience and effectiveness of the control strategies (e.g. the rate of positive tests, the number of positive cases that were contact traced, the temporal delay between the symptoms onset and the COVID-19 test outcome), and iii) the burden on the healthcare system (the hospital occupancy rate for COVID-19 cases, the number of intensive care units). High schools were closed on November 6th, and students were taught remotely; universities were encouraged to teach remotely.

Our data suggest that these measures effectively reduced the number of weekly contacts and the presence at work, and increased the adoption of personal protection measures. From the number of contacts participants specified by means of bins (e.g. “3-4”, “5-9”) (Q9 in SI Table S1), we average the lower bound of these bins (e.g. 3, 5) to account for the best case scenario. Figure 3A shows a drastic reduction in the estimated number of close contacts from outside the home for the different categories of respondents (students, employed, unemployed and retired). On November 10th, we observe an average reduction of 45% in the number of close contacts than those reported the week of October 13th. This decrease is particularly significant among students and the employed. By November 10th (one week after the colour-zones imposition), the reported number of close contacts was reduced by 81%, 35%, 65% and 55% for students employed, non-working and retired people, respectively, when compared to September. The reduction in the number of close contacts reported by students can be attributed to the restrictions on high schools and universities, which affected their daily number of contacts. When we average the number of close contacts in September and October versus the average number of close contacts from November to the end of December, respondents aged 18 to 20 years old limited their close contacts the most (−58%), followed by those aged 70 to 79 (−49%) and 21 to 29 (−46%) years old. This group, though, is the group with the smallest number of close contacts from outside the household. We refer to the SI Section S4 and Table S2 for additional details on the age range of participants. Employed respondents increased remote working by 9% on average, thus reducing their close contacts at work. We observe an increase both in unemployment (from 6% to 8%) and in those who reported being on unpaid leave (from 2% to 4%). We refer to the SI Table S5, Table S4, Table S6 and Table S7 for additional details.

**FIG. 3.**
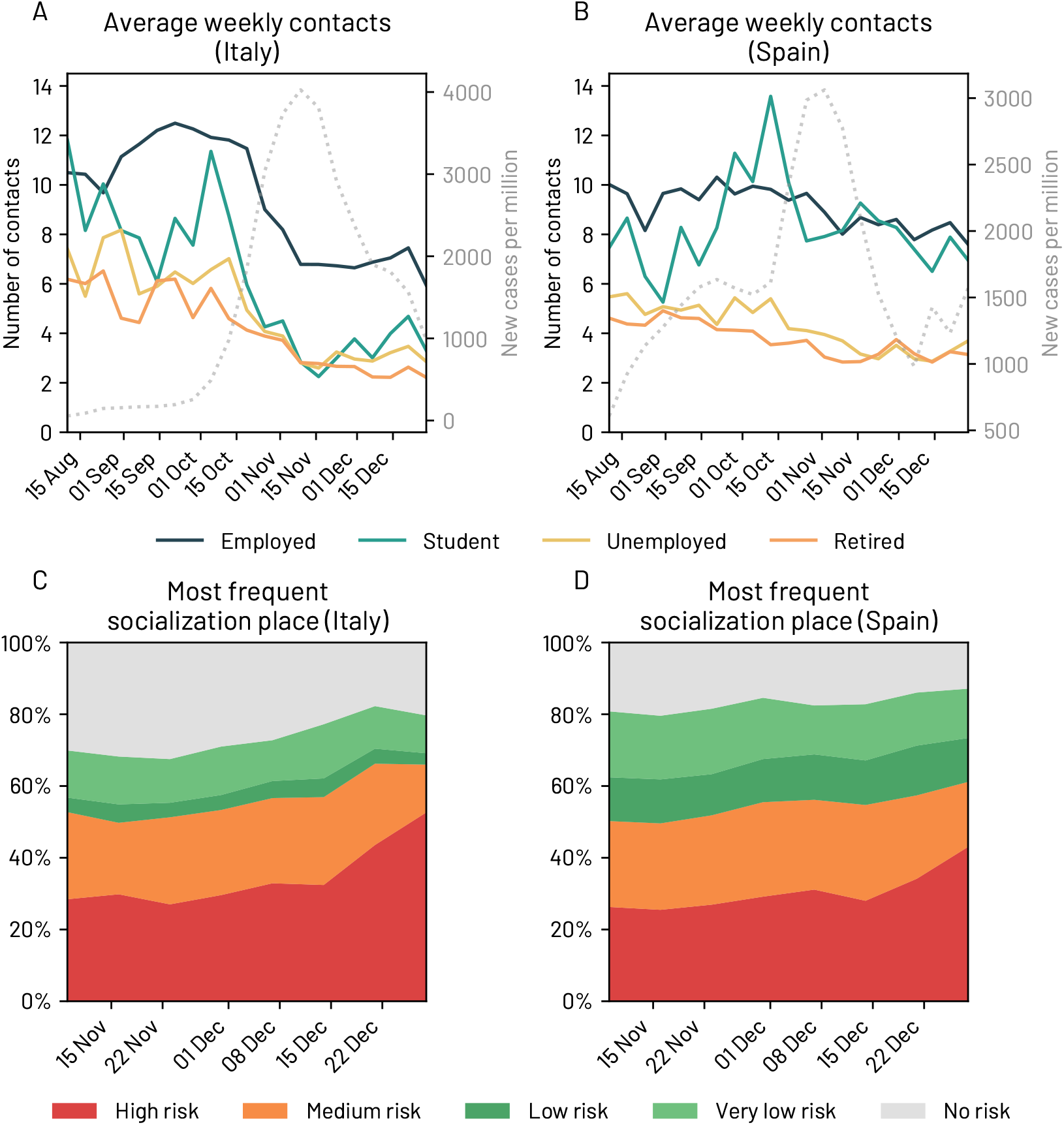
Average estimated number of weekly close contacts from outside the household together with the number of daily COVID-19 cases (dotted curve) in Italy A) and Spain B). C-D) Associated risk of the most frequent place of socialisation for Italy and Spain between November 1st 2020 and December 31st 2020.

Spain did not have a centralised COVID-19 mitigation strategy after the central government lifted the state of alarm on June 21st 2020 [31]. Hence, during the time period between June 21st and October 25th 2020, each of the 17 autonomous regions made their own decisions related to the pandemic, imposing restrictions as needed. However, in October 2020, all the autonomous regions in Spain (except for Madrid and the Basque Country) decided to adopt a common system to determine the pandemic risk via a set of indicators and implemented the same mitigation measures to slow the spread of COVID-19. They agreed to adopt a 5-level risk system ranging between *new normality (no risk)* to *extreme risk* based on their healthcare occupancy levels, the cumulative incidence, the positivity rate and the percentage of traced cases. Moreover, on October 25th, 2020, the government declared again a state of alarm [32], which enabled the declaration of a national curfew and gave power to local authorities to ban travel across regions, provinces and municipalities if needed. In terms of the measures, while every Autonomous Region decided to adopt their own, they were similar across regions and included limiting the mobility in the evening and at night, implementing regional/local confinements, reducing the maximum number of people in social meetings to 4-10 individuals, limiting the maximum capacity in locales to 25-50% and partially or totally closing shops in non-essential sectors, restaurants and bars. Together with the increase of confirmed COVID-19 cases, we find that these measures had a gradual and constant effect on the average number of close contacts participants reported having over time. For example, the average number of close contacts from outside the home for students and employed respondents decreased from more than ten in mid-September to around eight at the end of December. When we compare the average number of close contacts in the period between September and the end of October with those in the period between November and the end of December, respondents between 18 to 20 years old lowered their close contacts the most (−25%), followed by those aged 50 to 59 (−19%) and 21 to 29 (−19%) years old. We refer to the SI Section S4 and Table S2 for additional details. Surprisingly, teleworking decreased by 35%, from an average of 15% of teleworking in the new normality phase to 10% after the measures were implemented. The percentage of respondents reporting being on unpaid leave decreased by 52% (from 3% to 2%), while those who reported having lost their job rose by 29% (from 8% to 11%).

The increase in the number of COVID-19 confirmed cases, the measures implemented by the governments and the media probably contributed to a significant change in people’s perception of the risk associated with different places/activities and in the personal protection measures they adopted to minimise their risk of being infected with coronavirus. For example, in July 2020, over 60% of Italian respondents considered that going to the hairdresser entailed a low risk of getting a coronavirus infection. However, this percentage consistently decreased in the fall and winter of 2020, reaching 42% by the end of December 2020. Similarly, in Spain, the perception of safety when buying in small stores decreased by almost 20 percentage points in the same period. We also note a significant decrease of more than ten percentage points in both countries in the perception of how safe hospitals are, which could have negative consequences if many people chose not to go to the hospital when they should have gone for fear of getting infected with coronavirus. See SI Figure S11 and Figure S12 for additional information.

One of the primary goals of the governments’ pandemic mitigation strategies was limiting the amount of physical contact within the population. However, not all encounters are alike. In the survey, one question asked participants to report the type of context where they had more frequent socialisation with people outside of the household in the last week (Q10 in SI Table S1). We categorised the answers depending on their risk of infection, according to five levels of risk: *High risk* for indoor private environments (e.g. homes, private clubs) without adopting protection (HFS) measures; Restaurants and bars (indoors), schools and workplaces are considered to be of *Medium risk* as HFS measures are compulsory in such places; Restaurants and bars (outdoors) and other outdoor gathering activities would be labelled as *Low Risk* ; Nature, beaches and the street are considered to be *Very low risk* environments; and no social contacts outside of the household would be placed in a *No risk* category. Figure 3C and Figure 3D show the most frequent socialisation environment over time for Italy and Spain. We observe a gradual increase in the risk level of the main socialisation environment, as the percentage of those who report zero socialisation decreased and the percentage of people who reported social contacts in high-risk environments increased, probably due to the Christmas festivities in Italy and Spain. While the estimated number of weekly close contacts did not increase (Figure 3A-B), the types of environments where such contacts took place changed over time, with a significant increase in the prevalence of high-risk environments (Figure 3C and Figure 3D). See SI Figure S16 for a breakdown of the two countries.

### D. Perception of the Government Measures

As the number of confirmed COVID-19 cases rose again after July 2020, governments were forced to balance a new implementation of mitigation policies that would combine swift reactions and softer confinement measures to slow down the growth in the number of positive cases while minimising their economic and social impact. Thus, public health experts and policymakers needed to persuade their citizens to support and comply with the implemented measures.

Figure 4A shows how public opinion related to the implemented COVID-19 measures evolved over time in Italy and Spain (Q13). In Italy, as soon as the number of confirmed cases rose in the first week of October 2020, those who thought that the government should implement more measures increased from less than 40% to more than 50% around the first week of November. Consequently, the percentage of people thinking that the Italian policymakers had implemented enough measures fell to around 20%. Note that Italy implemented systematic pandemic containment regulation between the end of October and the first week of November. Surprisingly, in Spain, the percentage of respondents demanding more measures decreased in October 2020, even if the number of confirmed COVID-19 cases increased. Interestingly, the percentage of respondents demanding more measures only increased *after* the peak of infections in the second wave was reached in November 2020.

**FIG. 4.**
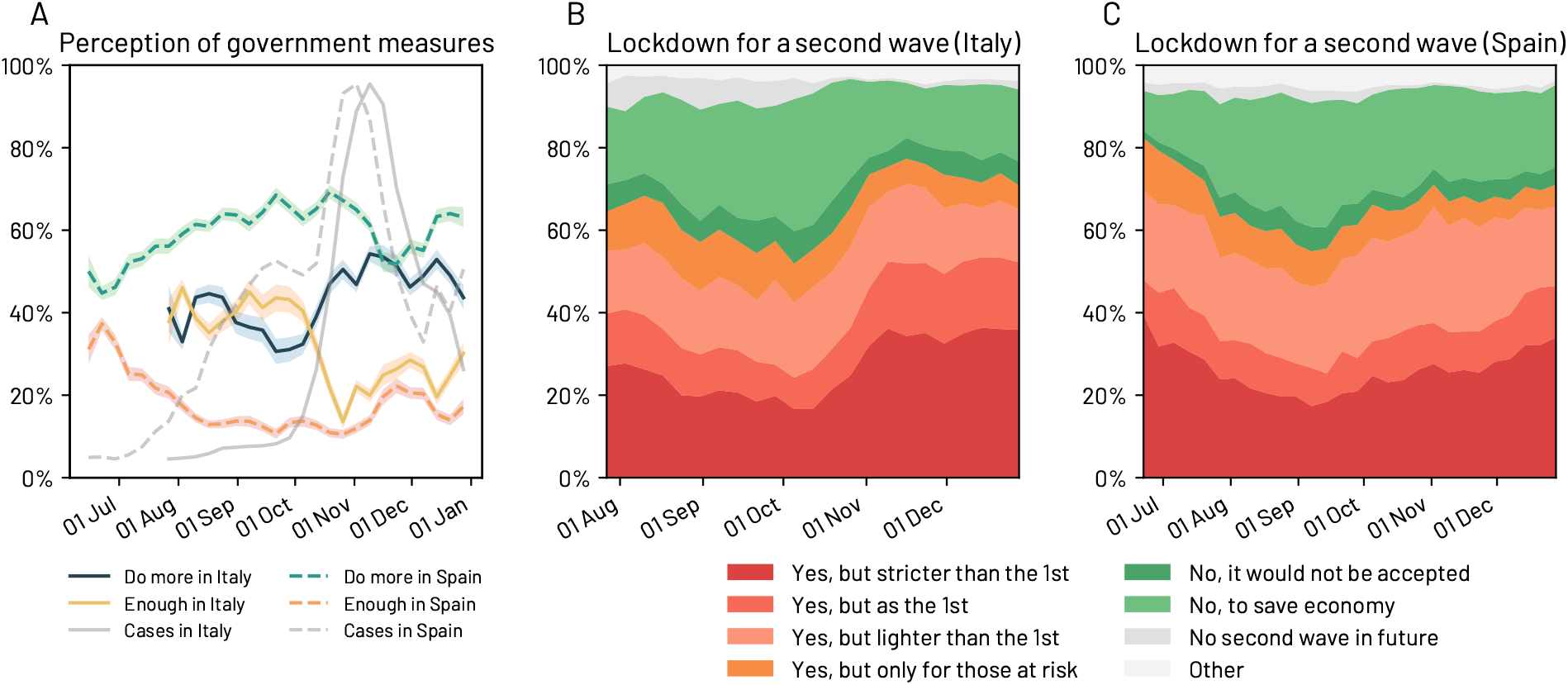
Participants’ perception of the Government measures and support for a potential lockdown change over time. In Italy (A-B) they seem influenced by the number of confirmed COVID-19 cases. We note that our analysis starts on July 31st and June 21st in Italy and Spain, respectively. In A) we report the 95% confidence intervals.

We also observe that the public opinion about a possible second lockdown changed over time in Italy and Spain (Q12 in SI Table S1). Both in Italy and Spain, we find an increase in the percentage of participants asking for a second lockdown as soon as the second wave hit the two countries in October 2020 (see Figure 4B and Figure 4C). Notably, in Italy, the percentage of respondents asking for a second lockdown increased by around 20% from the first week of October to the first week of November 2020; in Spain, the increase was over 20% between the first week of September until the end of the year.

## DISCUSSION AND IMPLICATIONS

In this paper, we have quantitatively analysed and compared the effectiveness of the TTI control strategies in Spain and Italy by analysing data collected using a large-scale online survey from June to December 2020. We have also studied the impact of the pandemic mitigation interventions on the respondents’ behaviour, including the number of close contacts, the context of socialisation, the personal protection measures that respondents reported to adopt to avoid COVID-19 infections, their perception of the safety of several activities and places, and their assessment of the government’s interventions to mitigate the pandemic. From our analyses, we draw several implications related to TTI and the mitigation interventions implemented to contain the spread of SARS-CoV-2.

### E. More efficient, holistic approaches to TTI are needed

During the coronavirus pandemic, many countries used a TTI control strategy to limit the number of infections and reduce the societal, economic and healthcare burden of COVID-19. Previous works have shown that TTI can in principle contain the spread of SARS-CoV-2 if physical distancing measures are kept [10–13, 33], although this control strategy is challenged by the role played by pre-symptomatic and asymptomatic transmission in the spread of coronavirus [34–38]. Moreover, TTI has been successfully used to prevent new outbreaks in countries such as South Korea [39], and Singapore [40]. However, the emergence of a second and third wave of COVID-19 cases in most European countries reveals how challenging it is to implement such a strategy effectively. In this paper, we have analysed how well Spain and Italy were able to trace, test and isolate their positive cases through the lens of an online, large-scale citizen survey.

In terms of *tracing*, we find that 29% of the close contacts of a positive coronavirus case were traced and 75% of the respondents who reported having tested positive when they filled the survey also reported having been traced. Moreover, we find that the types of contact that were mostly traced are different depending on the country. For example, Spain seemed to focus on tracing household members instead of tracing contacts from outside of the home. While tracing within the household is essential, the household members are likely to be already aware of their close contact with an infected individual in their own household. Moreover, tracing only the household members has a limited impact on keeping under control the transmission in workplace and community settings [41–43].

Hellewell *et al*. [11] investigated the success of contact tracing depending on the initial number of infections, the reproduction number, and the amount of pre-symptomatic or asymptomatic transmission. Their results show that most scenarios with a reproduction number of 1.5 were controllable, with roughly 50% of contacts successfully traced. However, scenarios with a reproduction number of 2.5 required the tracing of more than 70% of the contacts.

Our results show that the tracing coverage in Spain and Italy was far from these figures. Moreover, the two countries were especially weak in tracing people with the highest number of close contacts (50+). Our findings also suggest that as the COVID-19 incidence increased, the contact-tracing capacity decreased, as also observed by previous literature [44].

Given the limitations of manual contact tracing, various countries deployed digital contact tracing tools via a mobile app to complement the manual contact tracing efforts. Kretzschmar *et al*. [45] claim that digital contact tracing on its own would be more effective than conventional manual contact tracing alone even with only 20% of app adoption, due to its inherent speed. In the best-case scenario, digital contact tracing alone could reduce the reproduction number by 17%. More recently, several studies have shown the potential effectiveness of digital contact tracing using real-world contact patterns [43, 46] and in pilot studies in Switzerland, the United Kingdom (the Isle of Wight), and Spain (Gomera island) [47–49]. However, our data reveal the very limited role played by contact tracing apps. Despite having significant adoption figures in our sample (31% in Italy and 19% in Spain), only 1% of respondents who reported having had a close contact with an infected individual discovered it via the app. In addition, a small fraction of those got tested. This limited role played by digital contact tracing could result from a conjunction of factors, including technological limitations, low integration with local health policies, and delayed notifications. Thus, more detailed analyses on the real-world epidemiological effectiveness of the digital contact tracing apps would be needed [50].

Finally, we observe significant discrepancies between officially reported and our survey data, particularly in Italy. According to officially reported data in Italy, more than 86% of the positive cases would have been contact traced after November 6th 2020 (see SI Figure S17). According to our data, 75% of the contacts of infected individuals were called, but only 61% of infected participants reported being aware that their contacts had been called by contact tracers (see SI Figure S5). Although this difference might arise from a different definition of close contacts, citizen surveys represent an alternative source of data to be compared with officially reported figures.

The second pillar of the TTI strategy is *testing*. Several studies have emphasised the importance of minimising testing and tracing delays both in manual and digital contact tracing [11, 33, 45]. Kretzschmar *et al*. [45] show that a testing delay of more than one day requires a tracing delay no longer than one day and a tracing coverage of a least 80% of contacts to keep the effective reproduction number below 1. A testing delay of 3 days or longer would make it impossible for the most efficient tracing strategies to reach a reproduction number below 1. The authors conclude that reducing the testing delays –by shortening the time elapsed between symptom onset and a positive test result– would be the most crucial factor to increase the effectiveness of contact tracing, assuming the immediate isolation of all positive cases, which, as we show next, is not the case in practical terms. According to our data 41% of participants had to wait more than three days to receive their test result, while only 39% and 37% of Italians and Spanish, respectively, had the test results within one day. Opposition to get tested has been found to be a potentially significant barrier in the TTI strategy [51]. Although our data might be biased towards people willing to collaborate, we have not found evidence of a significant percentage of the population in Italy and Spain refusing to get tested (3% in Italy and Spain).

The final pillar in the TTI strategy is the effective *isolation* of all positive cases. According to our data, 54% of participants aged 18-59 years old would be unable to self-isolate if needed. While most respondents were unable to self-isolate due to sharing their home with others, we identified additional age and gender-dependent reasons: psychological reasons are the most frequent amongst the youth (20% and 16% of participants aged 18-29 years old in Italy and Spain, respectively); and having to take care of children is most likely reported amongst women aged 30 to 59 years old (28% of women vs 13% of men within the same age group in both countries). Previous work has highlighted the importance of providing isolation infrastructure as a critical COVID-19 control measure [22]. While Asian countries have provided hospitals and other facilities to support the isolation of all confirmed cases, European countries have only partly do it, mainly asking people to self-isolate in their homes, irrespective of their ability to do so [22, 52]. Planning the necessary infrastructure to support isolation, particularly to the youth and families with young children is of paramount importance [53].

### F. The importance of monitoring behaviour and perceptions

Our citizen survey allows us to compare the respondents’ self-reported behaviour before and after the pandemic containment strategies were deployed in Italy and Spain in the fall of 2020. Thus, through the lens of the survey, we can measure the impact of the government interventions on people’s behaviour, assess the compliance of the recommended individual protection measures, detect significant changes in behaviour that could lead to potential new waves of infections and estimate the population’s support to the government measures.

First of all, we find a very significant decrease in the estimated number of close contacts outside the home and a significant increase of teleworking and unemployed respondents after several containment measures (e.g. closures of museums, theatres, cinemas, gyms and swimming pools; restrictions to or closures of bars and restaurants; remote teaching activities in the majority of the high schools and universities; curfew) were implemented in Italy at the end of October of 2020. This result provides some supporting evidence to the effectiveness of these measures in limiting the number of close contacts among people and their potential in mitigating the spread of SARS-CoV-2. Coherent results were found in previous single-country [6] or multi-countries [5, 7, 54] data-driven studies as well as in previous modelling studies using fine-grained mobility data [55]. For example, Brauner *et al*. [5] analysed the effects of several government mitigation interventions on coronavirus transmission in 41 countries worldwide during the first wave of the pandemic. Their findings show that interventions such as the closure of school and universities, the ban of gatherings with more than ten people and the targeted closures of venues with high infection risk (e.g. bars, night-clubs, restaurants) were significantly more effective than other interventions (e.g. closure of all business venues, stay-home orders). Chang *et al*. [55] studied the hourly movements of 98 million people in the US and showed the high risk of specific venues (i.e. full-service restaurants, fitness centres, cafes, religious organisations) in terms of the number of potential close contacts and increase of infections. It is worth noticing that a more limited impact of the mitigation interventions was found in Spain, probably due to the de-centralised approach to implementing these containment strategies in each of the 17 Autonomous Regions.

In Italy, we find that students and unemployed respondents reduced their close contacts the most, reaching a 58% reduction in the youngest demographic group (18 to 20 years old). This result might contribute to explain both the psychological difficulties reported by the youth in terms of their ability to self-isolate and the reported negative psychological impact (e.g. anxiety, depressive symptoms) of the pandemic on students [56, 57].

Regarding the adoption of individual protection measures, hand washing and face mask-wearing were the most widely adopted measures by Italian and Spanish respondents. Conversely, physical distancing and ventilation of indoor spaces were the least adopted measures. Even though roughly 82% of our respondents reported wearing a mask as much as possible, masks might not be worn in the highest risk situations (e.g. private indoor spaces, with friends) and might be worn in lower risk, public environments, such as outdoors and streets. In fact, we find a noteworthy increase in the percentage of respondents who reported socialising in high-risk environments (i.e. indoors without protection measures) during the Christmas holidays, which might have been a side-effect of the closures of public gathering spaces, restaurants and bars during that time period. This increased socialisation in high-risk environments might explain the rise of the third wave of coronavirus infections in Spain after the Christmas holiday period.

We also find that the respondents’ demand for more government measures to combat the COVID-19 pandemic increased in Italy as the confirmed number of cases increased. However, in Spain, we do not identify such a pattern.

Finally, we show that Spanish respondents and women in both countries tend to be more compliant in adopting personal protection measures –such as wearing facial masks– and more conservative in their estimation of COVID-19 risk associated with different activities/places. The identified gender difference is aligned with previous studies on attitudes and behaviours during the COVID-19 pandemic [58] as well as with the literature showing that women tend to be more risk-averse and more open to government interventions than men [59, 60]. Overall, these results highlight the challenges posed by behavioural changes in response to new risks and the need for targeted risk communication strategies [58].

### G. Misalignment between the perception of coronavirus infection risk and real risk

From the survey answers, we observe that the perception of infection risk is heterogeneous and dependent on age and gender. Moreover, it changed significantly as the reported COVID-19 incidence increased. In July of 2020, over 41% of Italian respondents and 48% of Spanish respondents considered that one could go to a hospital with a low risk of getting a coronavirus infection. However, by December 2020, such percentage had dropped to 22% in Italy and 38% in Spain.

We also find a misalignment between the perception of risk and the actual risk. Some activities are considered to entail a higher risk of infection than those supported by actual data, whereas other activities are considered safer than they are. For example, during the study, flying by aeroplane is consistently considered the activity with the highest risk of getting a coronavirus infection (13% of Spanish respondents and 19% of Italian participants responded that one could fly by aeroplane with a low risk of infection). However, an aeroplane cabin is probably one of the most secure environments regarding the risk of a coronavirus infection [61]. Similarly, our data suggest that people consider beaches to be less safe than hairdresser salons and small stores, while indoor spaces would in principle be riskier than outdoor spaces, especially when social distancing measures are adopted [42, 62, 63]. Finally, people seem to perceive as less dangerous the activities or places that they are likely to do or go to. For example, young participants (aged 18-29 years old) considered practising sports less risky than older participants (aged 60+ years old). Similarly, more than 30% of young respondents (aged 18-29 years old) considered as safe to travel using public transportation, while only around 22% of older participants thought the same.

Based on these results, it would be essential to deploy public health communication campaigns to inform citizens about the actual safety of different activities and environments.

## CONCLUSION

During a pandemic, governments and societies must gauge real-time information about the effectiveness of non-pharmaceutical interventions, their impact on people’s behaviour and the population’s perceptions about them. However, deploying conventional surveys (e.g. pencil and paper surveys) in the midst of a pandemic has logistical and temporal challenges. Instead, online surveys allow a fast and cheap collection of data that might complement existing data sources and corroborate officially reported data.

The COVID19ImpactSurvey used in this paper was launched in March 2020 and has been regularly used by the Valencian Government [64] to support and evaluate their decision making. With over 600,000 answers to date, it is one of the largest online citizen surveys related to coronavirus. The answers to the survey have enabled the Government to understand better people’s behaviour, perception, and compliance with the confinement measures; the economic, labour and psychological impact of the pandemic; their ability to self-isolate and the efficacy of contact tracing. Similar efforts have been made to collect symptoms information [65–68] and behavioural data during the first wave [15, 69–71] of COVID-19. However, to the best of our knowledge, we are the first to collect such an extensive amount of data on self-reported human behaviour, and the control and mitigation strategies before and during the second wave of coronavirus infections in two countries.

Collecting a large sample of survey data (138,644 answers in two countries in our case) does not come without limitations. First, respondents have to be adults (at least 18 years old). Hence, students are only partially covered. Second, there might be a recall bias as respondents were asked about their behaviour, perceptions and situation in the last seven –sometimes fourteen– days (e.g. the number of close contacts in the last week). Third, there are self-selection and sampling biases [72] as the survey is filled out by volunteers who have learned about the survey via social media, WhatsApp, newspapers’ articles or Facebook ads, and who need to have access to a computing device (smartphone, tablet, PC) with an Internet connection. We mitigate these biases by applying weights to the raw data, so the resulting distribution closely matches the distribution of population-representative samples, and by deploying gender-balanced Facebook advertisement campaigns. Finally, note that our sample might have a different distribution of jobs than the population (see SI Figure S2).

In sum, citizen surveys are particularly valuable tools in situations of data scarcity where informed and timely decisions are needed. Online surveys also allow monitoring people perception and behaviour, enabling the design of more effective non-pharmaceutical interventions and better education and communication with the public. We plan to expand the survey to other regions, focusing on developing economies in Latin America.

## METHODOLOGY

### H. Data Collection and Processing

We analyse a subset of the answers to the COVID19ImpactSurvey, an extensive, anonymous, online citizen survey about COVID-19 [15]. Launched on March, 28th 2020, in Spain, the survey has since then collected over 600,000 anonymous answers from 11 countries, and it is available online at https://covid19impactsurvey.org/. Participants must declare being 18 years or older to be able to fill the survey.

The project has been approved by the Responsible Research Board at the University Miguel Hernandez. The Board is composed of the following members: Alberto Pastor, Tomas de Domingo, Javier Saez, Eugenio Vilanova, Vicente Micol, Antonio Guerrero and Ana Maria Madariaga

The survey consists of 26 questions which collect information about the participants’ demographic and household situation (Q1-Q6); their social contact behaviour (Q9-Q11); their support for the government measures deployed to contain the spread of COVID-19 (Q12) and for a potential lockdown (Q13); the economic impact of the pandemic in their lives (Q17-Q18); their ability to self-isolate if needed (Q19); the individual protection measures that they adopt to protect themselves against COVID-19 infections (Q20); whether they have been tested (Q24) for COVID-19; their willingness to get tested for COVID-19 infections (Q25) and the psychological impact of the pandemic in their households (Q26). We describe in SI Table S1 all the questions.

We analyse the data from Spain and Italy for the time period between June 2020 and December 2020. SI Table S3 and Figure S1 summarise the statistics of the data.

Survey answers were collected from volunteer respondents who learned about the survey via social media channels, word-of-mouth, universities and news organisations. We used Facebook and Instagram ads as an additional channel to recruit volunteers. This approach gave us a straightforward method to obtain a broad sample of users across each country. We used the same ads, images and budgets in both countries. We did not use any targeting feature except for gender, where we used separate budgets to balance the numbers of male and female respondents. The cost-per-successful-response was €0.24 and €0.11 for men and women respectively in Spain, and €0.13 and €0.07 for men and women respectively in Italy. Most of the cost difference between men and women could be explained by their higher willingness to click on ads, where women clicked on 2.1% of ads shown vs 1.3% of men. Both men and women had similar completion rates (54%) once they began answering the survey.

To further validate our methodology, in April 2020 we commissioned an IPSOS.digital FastFacts panel of a cohort of 1,000 representative general population respondents aged 18-65 in Spain. This validation was done after we started using Facebook ads, but before the time period covered by this paper. The results of the IPSOS panel were generally within the margin of error of our survey results for the same time period and ages.

### I. Selection of the time periods

We divided our analysis into two phases: (1) Phase I -new normality and (2) Phase II -second wave. In Italy, the Phase I period starts on June 15th, 2020 when there was a relaxing of many of the implemented confinement measures, including cinemas, theatres and discos [73]. However, we started our survey in Italy on July 31st. Phase I ends on October, 26th 2020 with the deployment of the first mitigation measures to reduce the spread of COVID-19 [27] and the subsequent national colour system imposed to restrict movements and social gatherings [30]. In Spain, Phase I starts on June, 21st 2020 with the “new normality” phase, when most of the confinement measures were lifted and minor restrictions, such as defining a maximum occupancy in shops are handled by each autonomous community independently [31]. It ends on October, 25th 2020 with the re-deployment of a state of emergency across the country [32].

## Data Availability

The data that support the findings of this study are available on request from the corresponding author M.D.N.

## AUTHOR CONTRIBUTIONS

All authors conceived the study. K.R. and N.O. designed and launched the COVID19ImpactSurvey. M.D.N., K.R. and N.O. conducted the analyses and analysed the results. All authors wrote and reviewed the manuscript.

## DATA AVAILABILITY

The data that support the findings of this study are available on request from the corresponding author M.D.N. Kontis, V. *et al*. Magnitude, demographics and dynamics of the effect of the first wave of the covid-19 pandemic on all-cause mortality in 21 industrialized countries. *Nature Medicine* **26**, 1919–1928 (2020).

## Footnotes

a https://www.gov.uk/government/news/new-campaign-to-prevent-spread-of-coronavirus-indoors-this-winter

