## Supplementary Information for "The Impact of Control and Mitigation Strategies during the Second Wave of COVID-19 Infections in Spain and Italy"

#### CONTENTS

S1. Survey questions 2

S2. Additional figures 6

S3. Waiting time to obtain the test results 11

S4. Perception of COVID-19 risk in places and activities 12

S5. Additional tables 21

### S1. SURVEY QUESTIONS

Table S1 depicts the questions whose answers have been analysed in this paper. The survey has a total of 26 questions. However, there are a few conditional questions such that the number of the questions is that a person might answer could be larger than 26.

| Question | Possible answers |
|---|---|
| **Demographic and Household information** | |
| Q1. What is your age range? | [18-20; 21-29; 30-39; 40-49; 50-59; 60-69; 70-79; 80+] |
| Q2. What is your gender? | [Male; female; another gender] |
| Q3. Postal code/Zip code | Text entry |
| Q4. Type of home | [Single family house; apartment/flat; shared apartment/flat; other shared accommodation; other] |
| Q5. Number of people in the home (including you) | [1; 2; 3; 4; 5+] |
| Q6. Age(s) of people in the home (excluding you, check all that apply) | [10 or less; 11-20; 21-29; 30-39; 40-49; 50-59; 60-69; 70-79; 80+] |
| **Tracing** | |
| Q9. In the last seven days, approximately how many different people that live outside your home have you had close contact with? (meaning more than 15 minutes and a distance closer than 2 meters) | [No one; 1-2; 3-4; 5-9; 10-19; 20-29; 50+] |
| Q9_1. (only for those who had close contact with someone from outside the home) What was your most frequent type of close contact in the last seven days from outside your home? [We added this question on October 10, 2020] | [coworker; client; family/friends; school/university; concert/show/disco/club/conference; shopping; as a customer (bank, office); doctor/clinic/hospital; travel (including public transportation)] |

| | |
|---|---|
| Q10. In the last seven days, what was the most common location that you spend time socializing with your friends, relatives and acquaintances that live outside your home? [We added this question on November 12, 2020] | [Private house, apartment, residence or club while being strict about masks, distancing and ventilation; Private house, apartment, residence or club while more relaxed about masks, distancing and ventilation; Restaurant/coffee shop/bar/disco (indoors); Restaurant/coffee shop/bar/disco (outdoors); workplace; School/university; on the street, park, other public space; At the beach; in nature outside the city; other location; I did not socialize with anyone that lives outside my household] |
| Q11. Have you had physical contact with someone diagnosed with coronavirus (in the last seven days)? (check all that apply) | [Member of household; Family outside household; friend or acquaintance; coworker; Cleaning staff/caretaker/etc; Patient (in case of medical staff); client/customer; student; Unknown person (I was notified by the app or a contact tracer); None that I know of] |
| Q11_1. (only for those who respond that they had had a close contact with an infected individual) Have you been contacted by a doctor or a contact tracer about your close contact with someone infected with coronavirus? [We added this question on August 10, 2020] | [Yes; No; No, but I was notified by the app] |
| Q24_3. (only for those who tested positive) Did your doctor or health authority ask you to identify your close contacts in order to trace the infection? [We added this question on July 24, 2020] | [No, I was not asked about my close contacts; Yes, but none of my close contacts were called (to my knowledge); Yes, and some of my close contacts were called; Yes, and some of my close contacts were called and tested for coronavirus] |
| **Testing** | |
| Q24. Have you been tested for coronavirus? | [a) Yes, I am waiting for my result; b) Yes, the test said that I have coronavirus; c) Yes, the test said that I had coronavirus, but I am now recovered; d) Yes, the test said I do not have coronavirus (recently); e) Yes, the test said I did not have coronavirus (more than one month ago); f) No; I prefer not to answer] |

| | |
|---|---|
| Q24_1.(only for those who respond having been tested) In total, how long did it take (or are still waiting) to get an appointment, get tested for coronavirus and receive the results? [We added this question on September 25, 2020] | [1 day; 2 days; 3-4 days; 5-7 days; 8-13 days] |
| Q25. Are you currently trying to get tested for coronavirus due to having symptoms or having been recently exposed to an infected individual? | [Yes, I have an appointment to get tested and I am waiting for it; Yes, I am trying to get an appointment to get tested; Yes, but there are no tests available; Yes, but I do not know how to get tested; Yes, but the test is too expensive; No, but I want to take it; No, I do not think I need it; No, and I would refuse to take it; I prefer not to answer] |
| **Isolating** | |
| Q19. If you were diagnosed with coronavirus and had to be quarantined for at least 2 weeks, would you be in any of the following situations? (check all that apply) | [I could not isolate myself from other people in my home; I would have to continue taking care of other people (children, parents...); I depend on a caregiver; It would be difficult for me to get medical leave from work; I could lose my job; I could not afford it financially; It would be impossible for me psychologically; I would be afraid of discrimination or stigmatization; None of the above] |
| **Behavior and Perception** | |
| Q12. Do you think there should be a another lockdown if there is another wave of coronavirus? | [Yes, similar to the first lockdown; Yes, but stricter than the first lockdown; Yes, but less strict than the first lockdown; Yes, but only for people who are at risk; No, the economic and/or social cost would be too high; No, it would not be accepted by the population; I do not think there will be another wave; None of the above] |
| Q13. Do you believe that the measures the government has taken are enough to contain the spread of coronavirus? | [No, but should be stricter; Yes, are about right; Yes, but are too strict; Prefer not to respond; I do not know] |

| | |
|---|---|
| Q15. Which of the following activities do you think can be done with a low risk of coronavirus infection? (check all that apply) | [Practicing individual sports; Having friends visit you at home; Attending religious services with limited seating; Attending school like in some European countries; Going to small businesses with appointment (hairdresser, etc); Going to small shops while maintaining a safe distance; Having drinks at a bar on an open terrace with a group of people; Going to restaurants with limited seating; Receiving treatment at a hospital; Taking public transportation with space between seating; Going to the beach; Traveling by air; None of the above] |
| Q20. Do you take any of the following measures to prevent the transmission of the coronavirus? (check all that apply) | [I wear a mask as much as possible; I avoid crowded situations; I do not shake hands, give hugs or kisses to anyone who live outside my home; I regularly disinfect/wash my hands; I keep my physical distance of at least 1.5 meters (6 feet) from others; I limit the number of people that I am in close contact with; When indoors, I make sure there is good ventilation; I have installed my government's contact tracing app on my phone; None of the above] |
| Q21. Do you take any of the following measures to prevent the transmission of the coronavirus? (check all that apply) | [I wear a mask as much as possible; I avoid crowded situations; I don't shake hands, give hugs or kisses to anyone who live outside my home; I regularly disinfect/wash my hands; I keep my physical distance of at least 1.5 meters (6 feet) from others; I limit the number of people that I am in close contact with When indoors; I make sure there is good ventilation; I have installed my government's contact tracing app on my phone; I would be willing to get vaccinated immediately when the coronavirus vaccine is available; None of the above] |
| **Tele-work** | |
| Q17. Have you worked any time since March 1st, 2020? (before the beginning of the coronavirus crisis) | [Yes; No; No, I am a student; No, I am retired] |
| Q17_1 (only for those who respond that they have worked) Have you gone to work in the last seven days? | [Yes; Yes, with reduced hours; I am on leave or teleworking because I am in quarantine due to coronavirus; No, but I am teleworking; No, I am on paid leave (vacation, maternity, etc); No, I am on unpaid leave; No, I have lost my job or stopped working] |

| | |
|---|---|
| Q18. What is your main job? | [Essential services (police, firefighter, medical personnel); Retail; Manufacturing; Health and social services; Hospitality (restaurants, bars, hotels, etc); Education; Government or defense; Construction; Transport; Administrative assistant and similar; Professional, technical or scientific services; Farming, fishing or other food production; Press or communication; Household employee; Financial; Artist, recreation and entertainment; Sanitation, cleaning, garbage collection; Other services] |

TABLE S1: COVID19ImpactSurvey questions analyzed in this study.

### S2. ADDITIONAL FIGURES

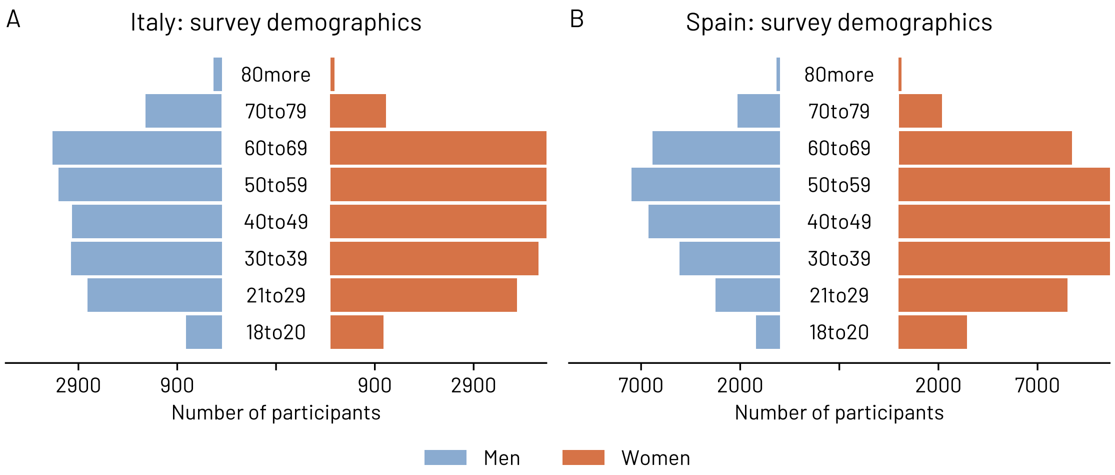


FIG. S1. Demographic composition of the respondents in Italy and Spain.

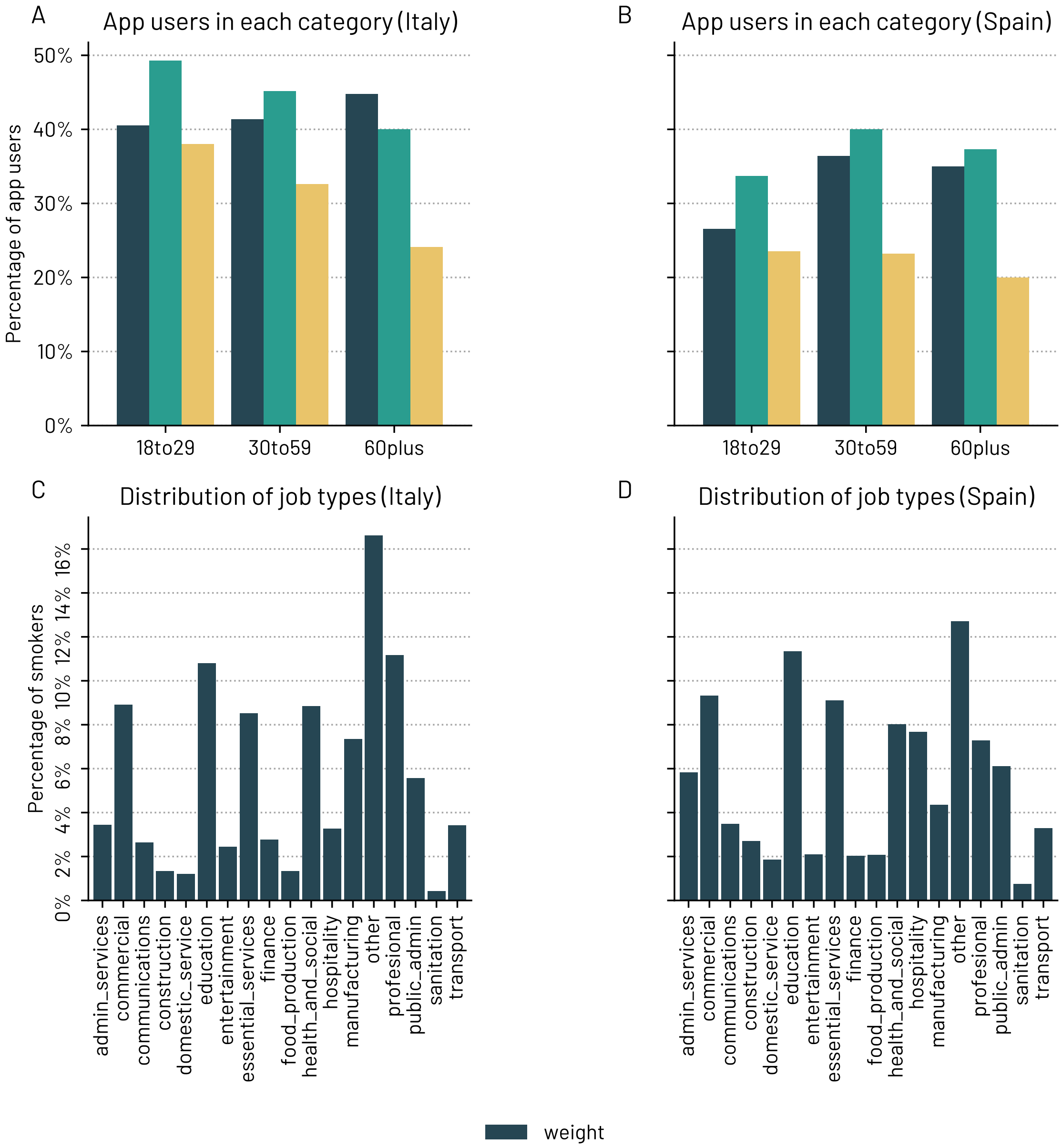


FIG. S2. Distribution of app users (A-B) and job types (C-D) in the respondents per age group in Italy and Spain.

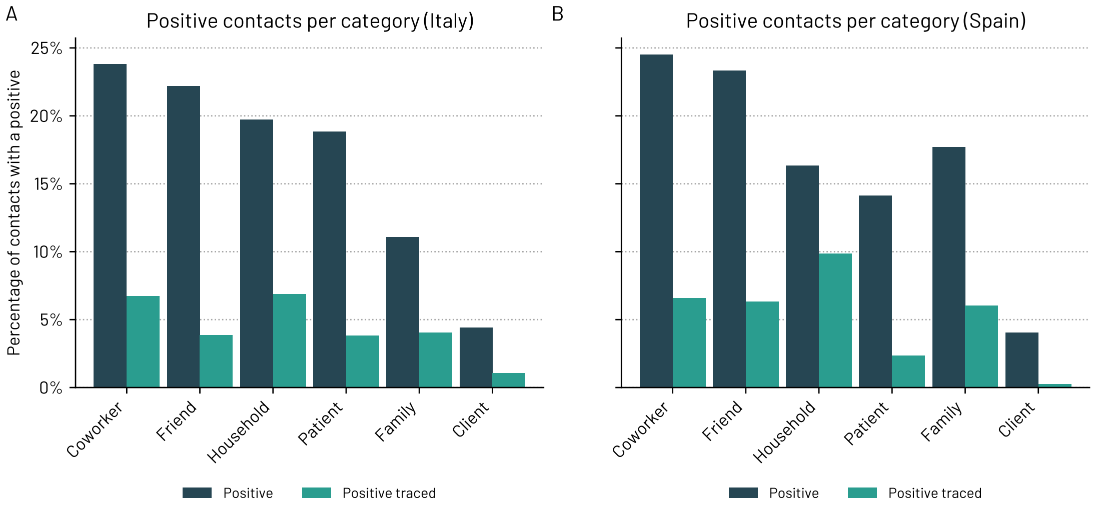


FIG. S3. A-B) The vast majority of respondents who reported having had a close contact with a positive case were not traced nor contacted during the Phase I - *new normality*. A person is traced if the doctor or the health authority identified his/her contacts. A person is contacted when some of the close contacts were called by the doctor or the health authority. C-D) Percentage of contacts with a positive and positive traced by contact type.

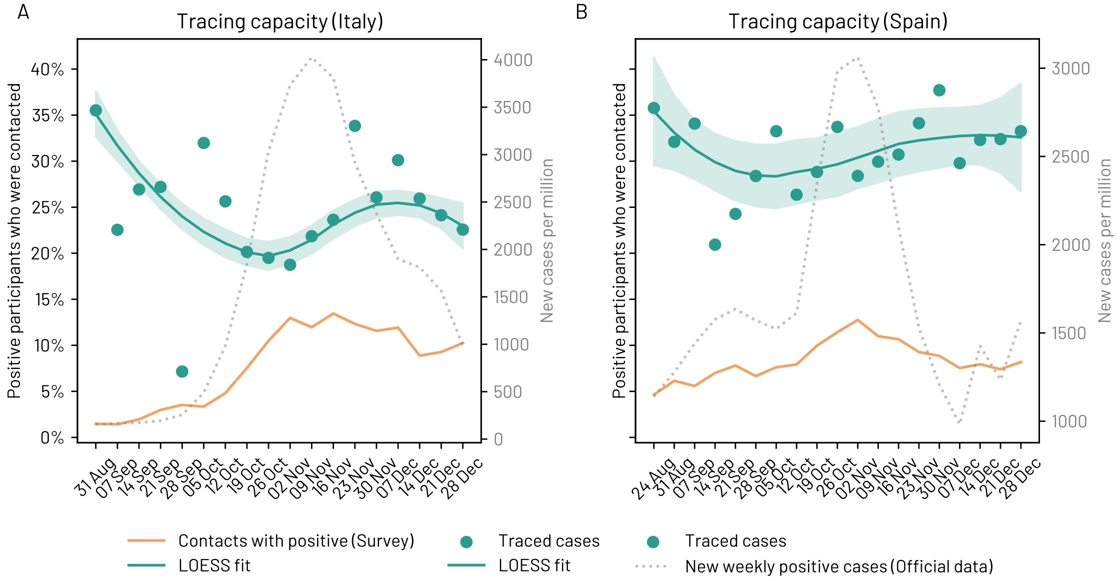


FIG. S4. Tracing capacity over time and daily number of COVID-19 positive cases. We report the 95% confidence intervals.

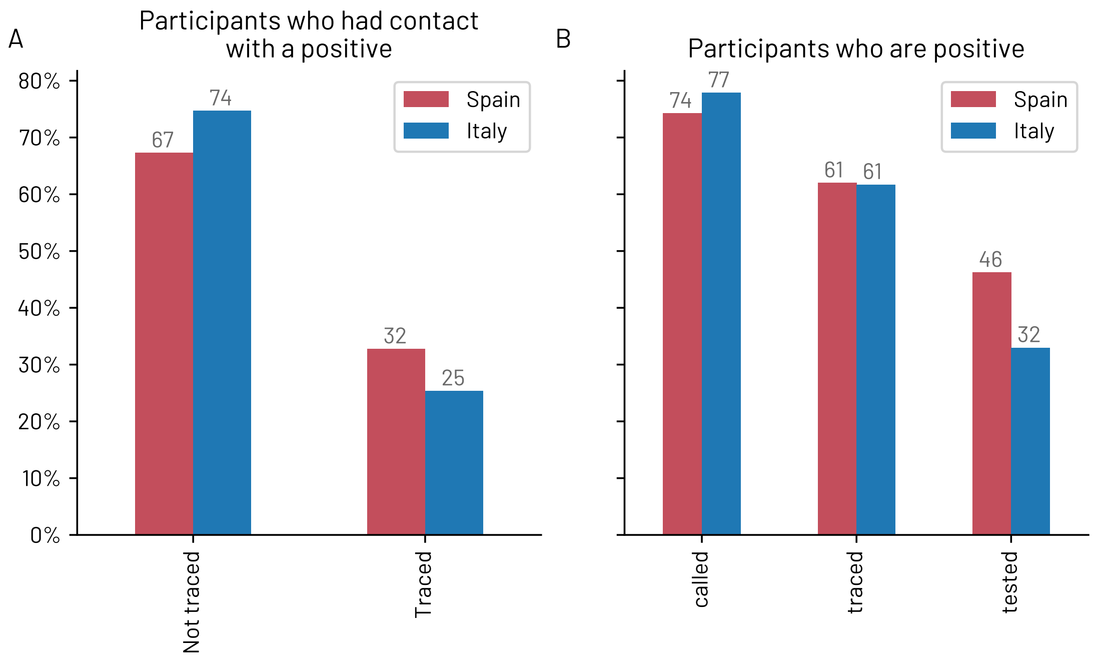


FIG. S5. Tracing statistics during the Phase I - *new normality.* A) Percentage of participants who reported having had a close contact with a positive case and having been contacted by the health authority. B) Percentage of respondents who reported testing positive when they filled the survey and having been contacted by the health authority.

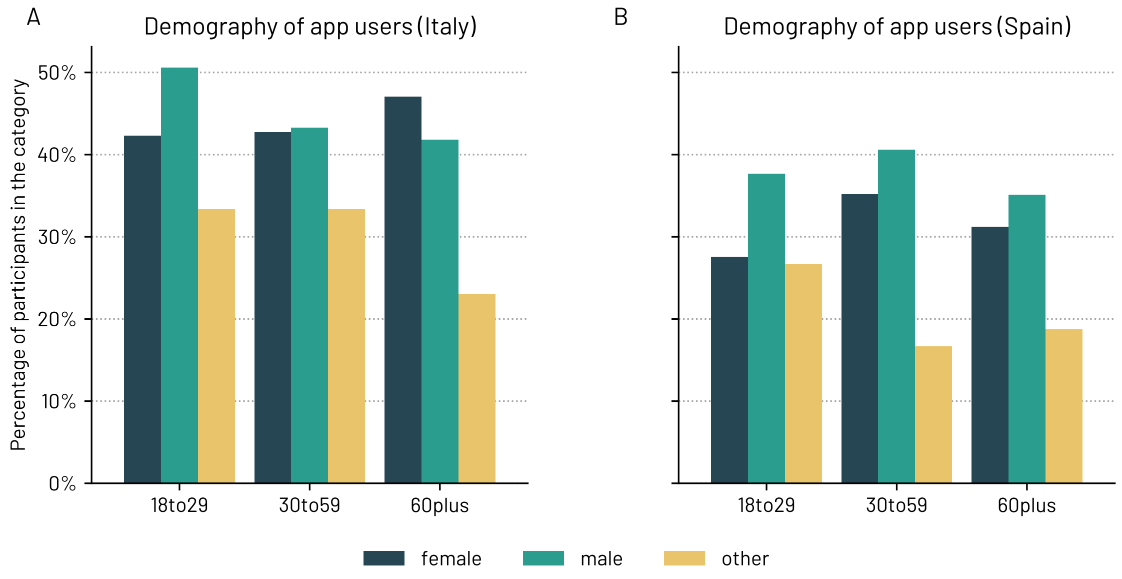


FIG. S6. Percentage of app users per each demographic category in A) Italy and B) Spain during the Phase I - *new normality.*

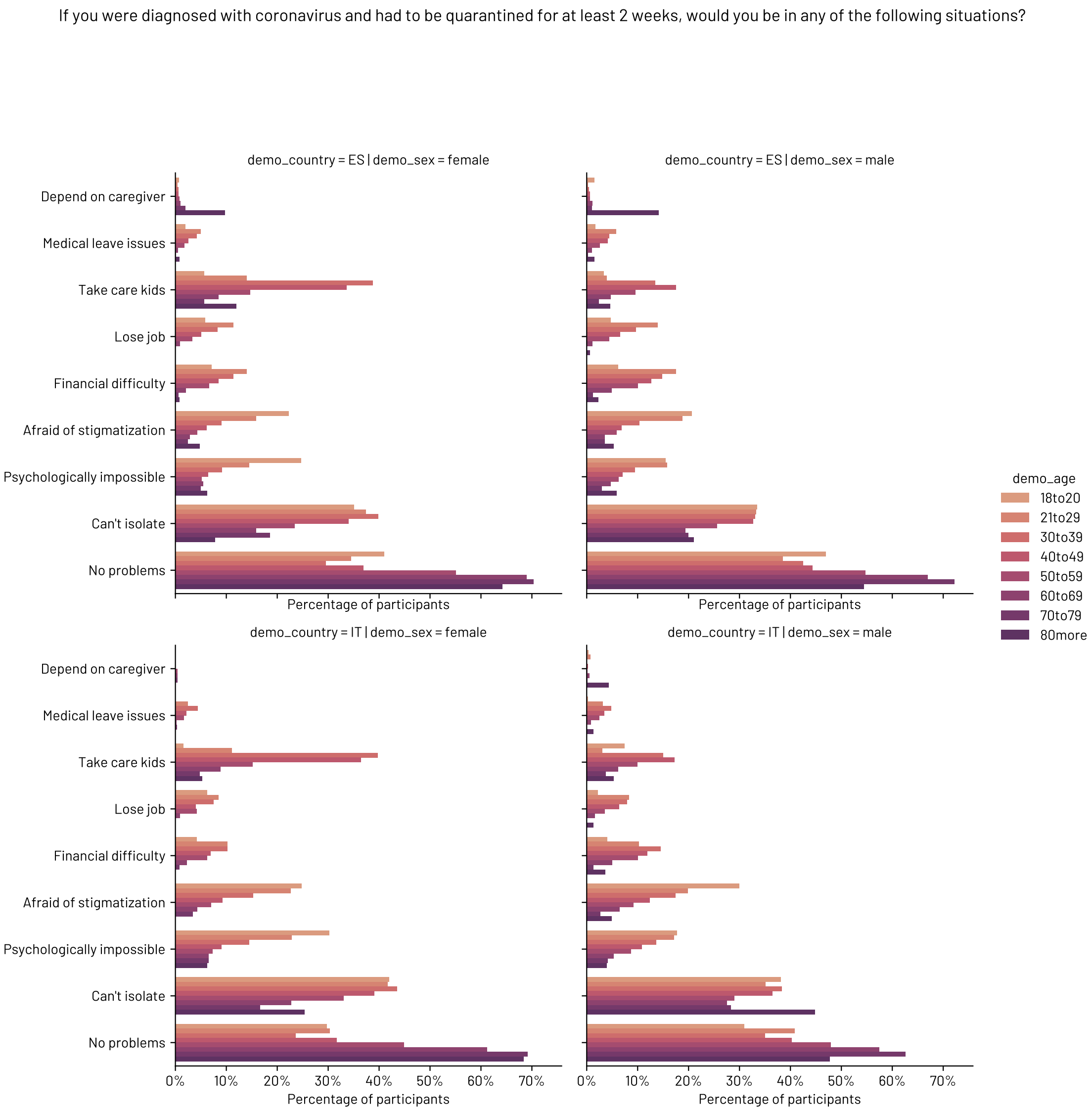


FIG. S7. Reported barriers in the case of being diagnosed with coronavirus and having to be quarantined for at least 2 weeks during the Phase I - *new normality*.

### S3. WAITING TIME TO OBTAIN THE TEST RESULTS

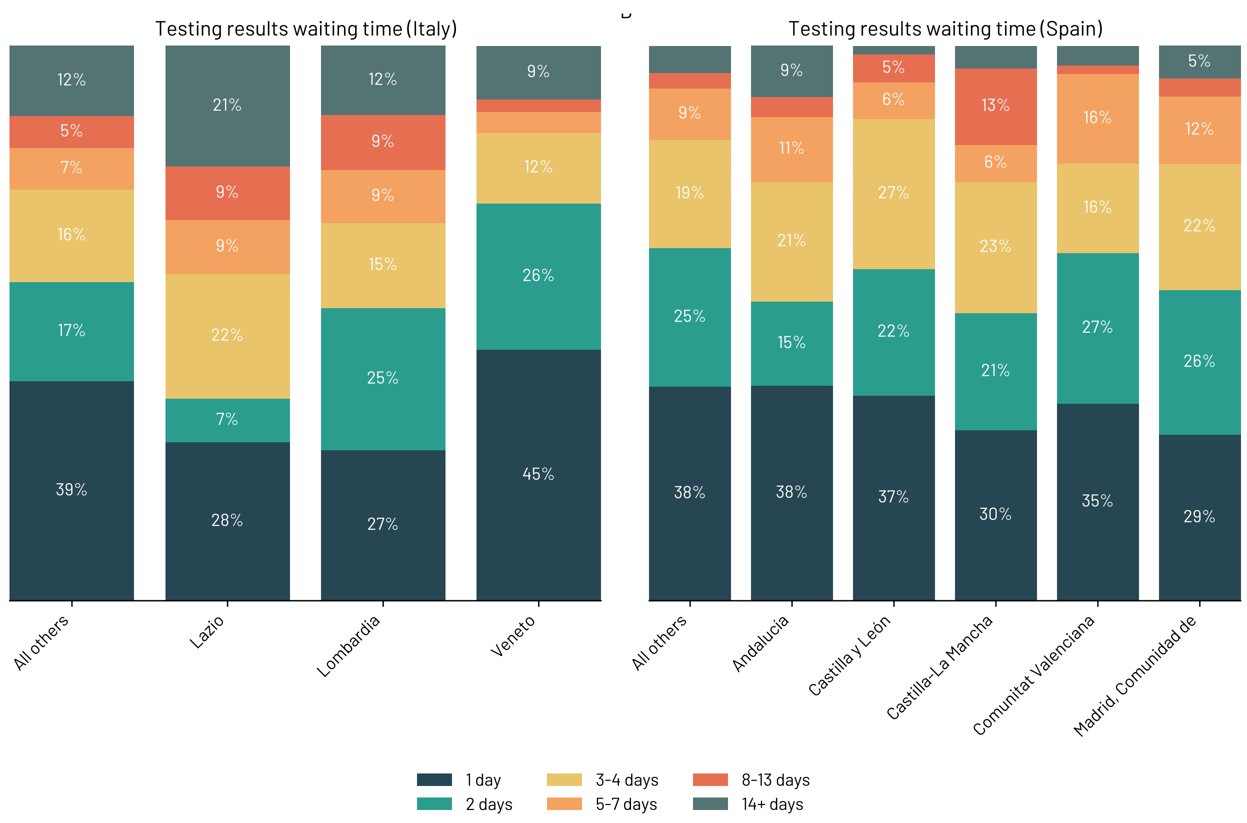


FIG. S8. Time to get the test results for each region in Italy (A) and Spain (B).

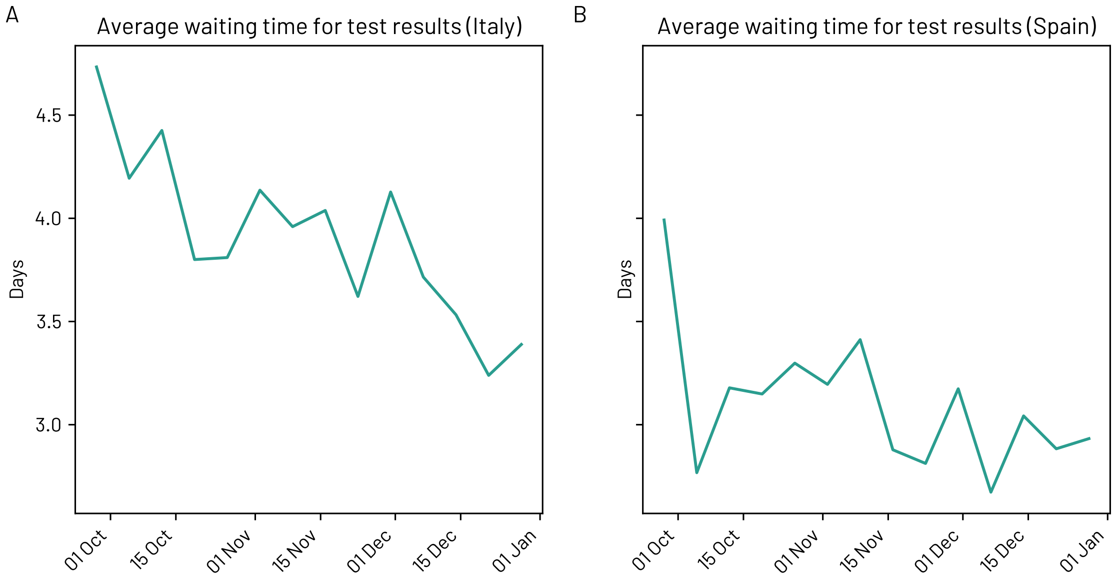


FIG. S9. Time to get a test results, over time, for Italy and Spain during the Phase I - *new normality*.

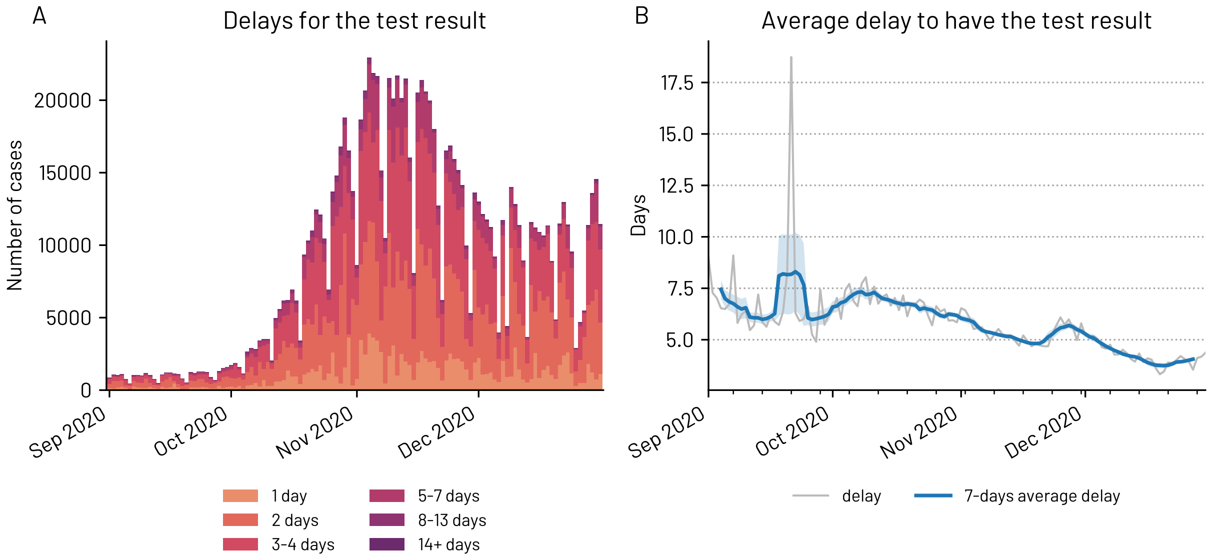


FIG. S10. Italian delays to get the COVID-19 result. Official data from the ISS. In B) we report the 95% confidence intervals.

### S4. PERCEPTION OF COVID-19 RISK IN PLACES AND ACTIVITIES

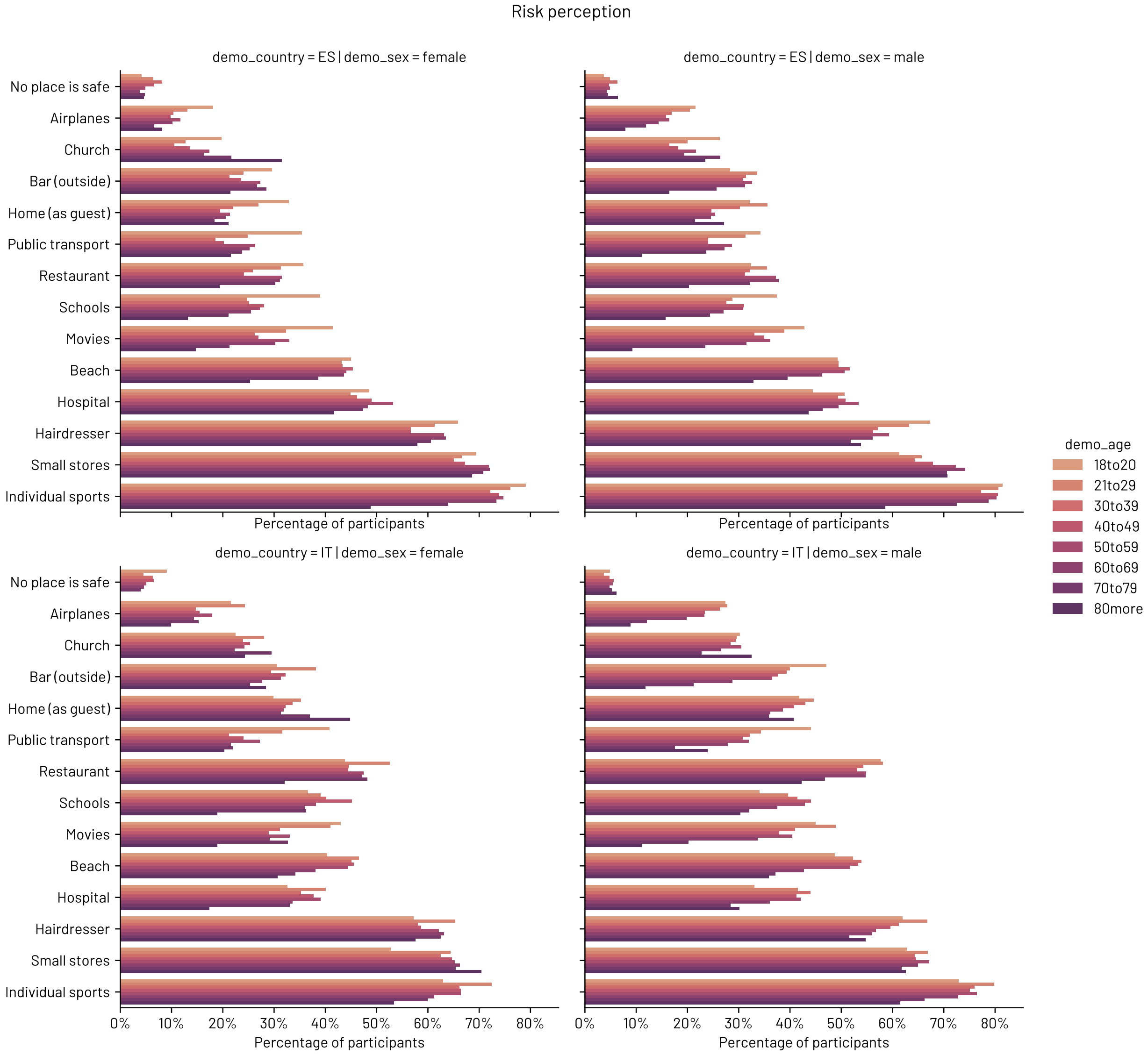


FIG. S11. Perception of safety per place, country, and demographic group during the Phase I - *new normality*.

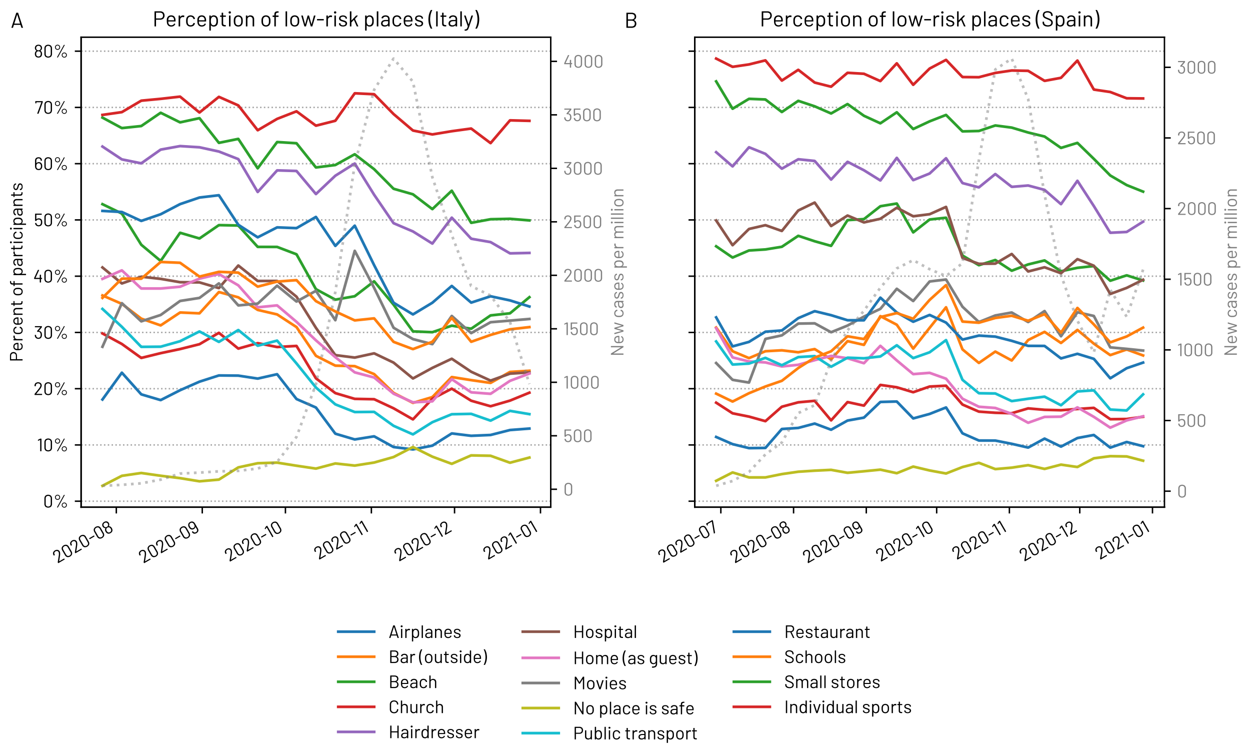


FIG. S12. Evolution of the perception of safety of activities and places over time.

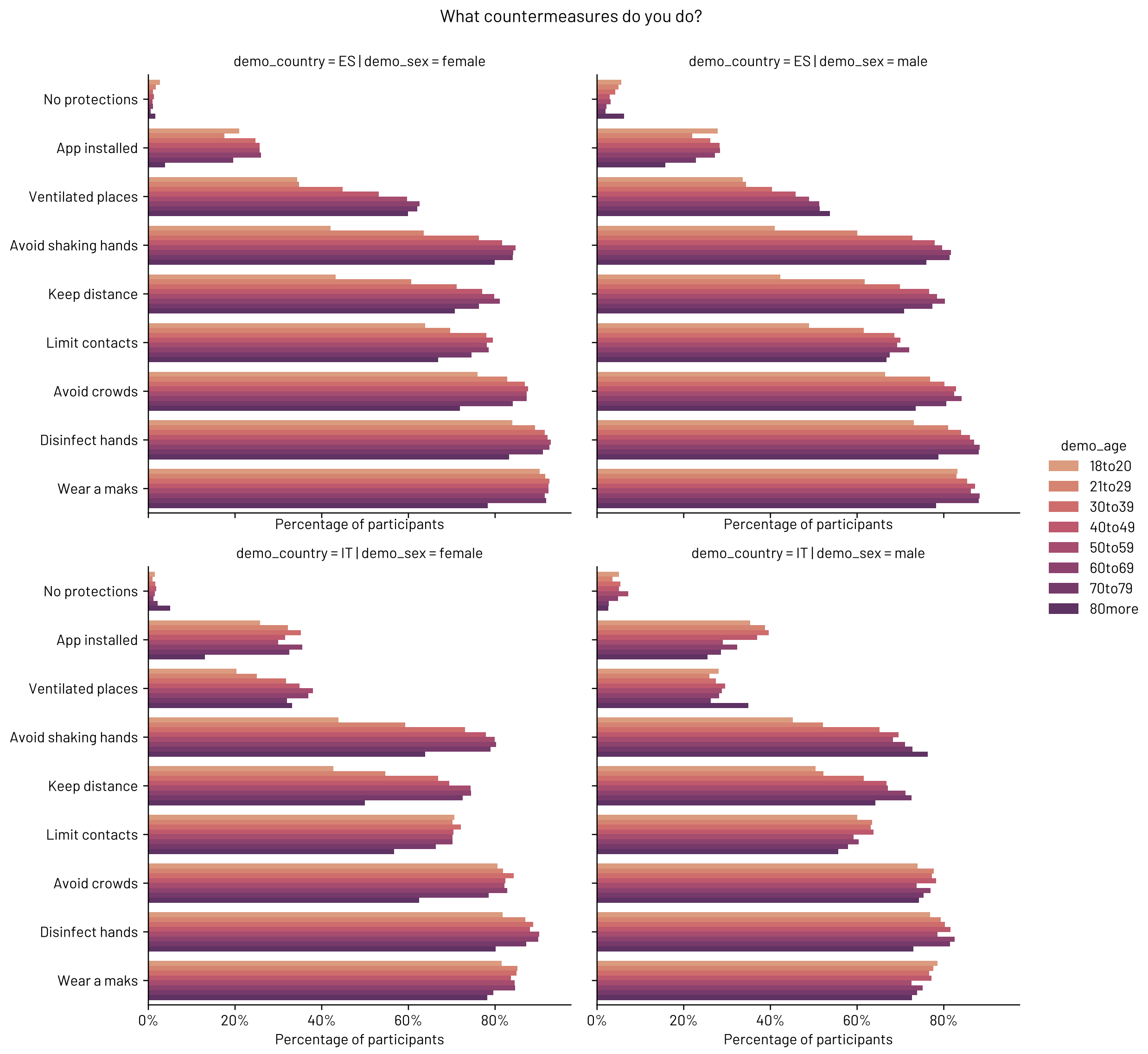


FIG. S13. Individual protection measures per country and demographic group during the Phase I - *new normality*.

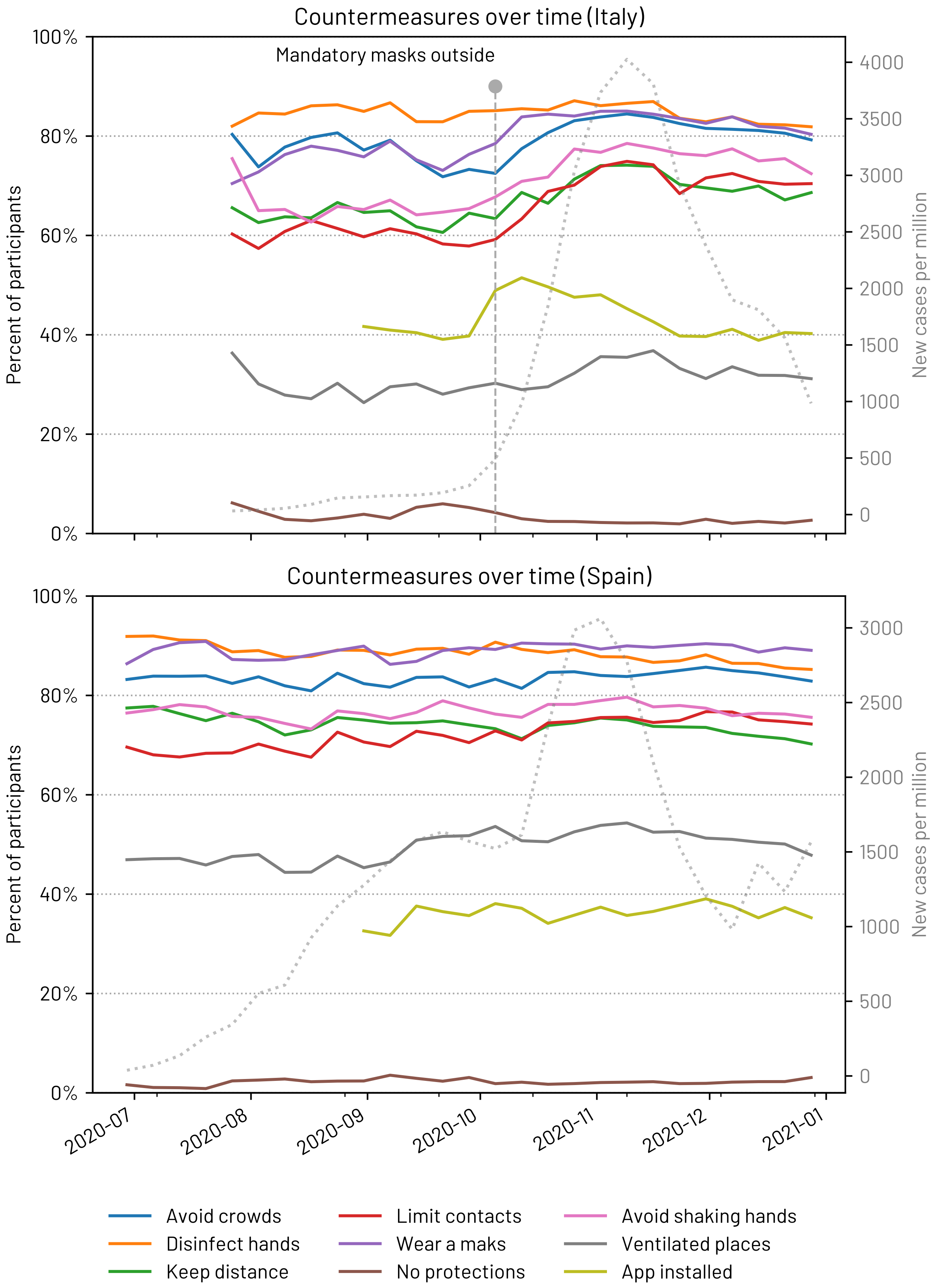


FIG. S14. Evolution of the adoption of individual protection measures over time.

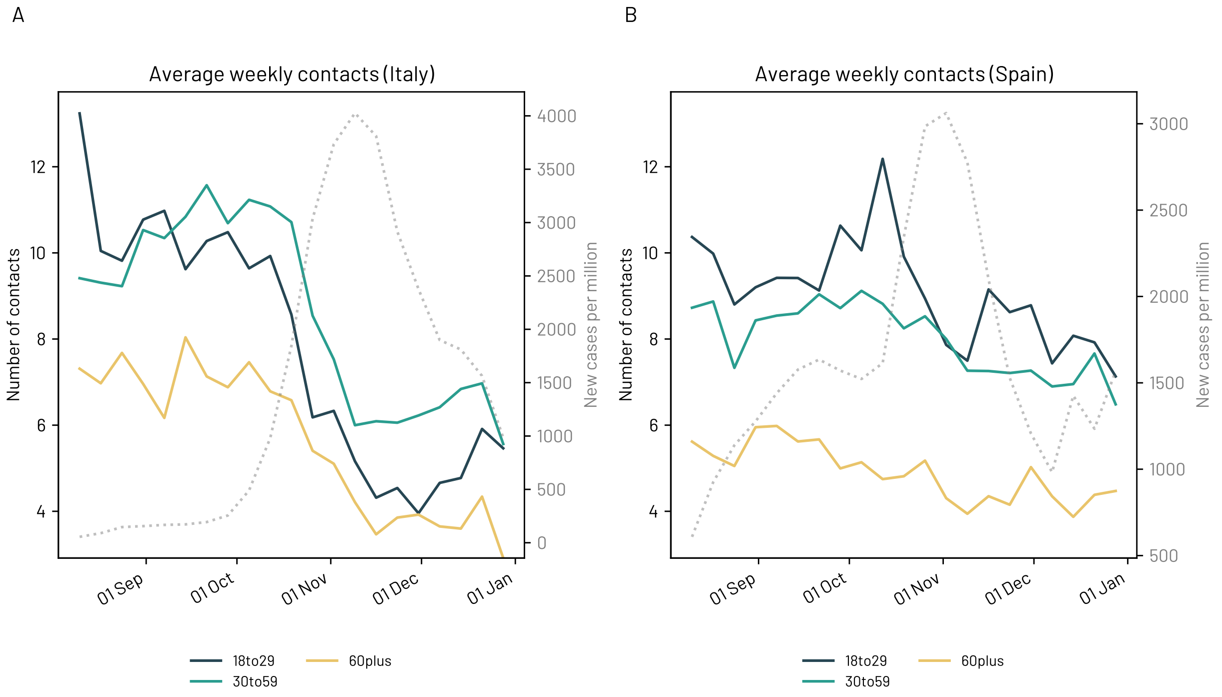


FIG. S15. Average number of weekly close contacts from outside the household per age range (18 to 29, 30 to 59, and 60+ years) in Italy and Spain.

| Country | Age range | Period I | Period II | Percentage difference |
|---|---|---|---|---|
| Spain | 18to20 | 11.52 | 8.63 | -25.15 |
| | 21to29 | 9.07 | 7.67 | -15.49 |
| | 30to39 | 8.96 | 7.28 | -18.71 |
| | 40to49 | 8.62 | 7.32 | -15.10 |
| | 50to59 | 8.39 | 6.77 | -19.33 |
| | 60to69 | 5.58 | 4.58 | -17.96 |
| | 70to79 | 3.76 | 3.11 | -17.50 |
| | 80more | 5.43 | 6.96 | 28.04 |
| Italy | 18to20 | 10.31 | 4.34 | -57.88 |
| | 21to29 | 9.90 | 5.39 | -45.56 |
| | 30to39 | 11.22 | 6.58 | -41.33 |
| | 40to49 | 11.06 | 6.51 | -41.17 |
| | 50to59 | 10.66 | 6.78 | -36.43 |
| | 60to69 | 7.72 | 4.45 | -42.34 |
| | 70to79 | 5.35 | 2.73 | -48.95 |
| | 80more | 5.90 | 3.81 | -35.52 |

TABLE S2. Percentage of reduction in the number of close contacts from outside the household between Period I (new normality) and Period II (second wave). We refer with the former with the period before the October 26th in Italy and the November 1st in Spain. We refer with the latter with the period of time after Period I and the end of 2020.

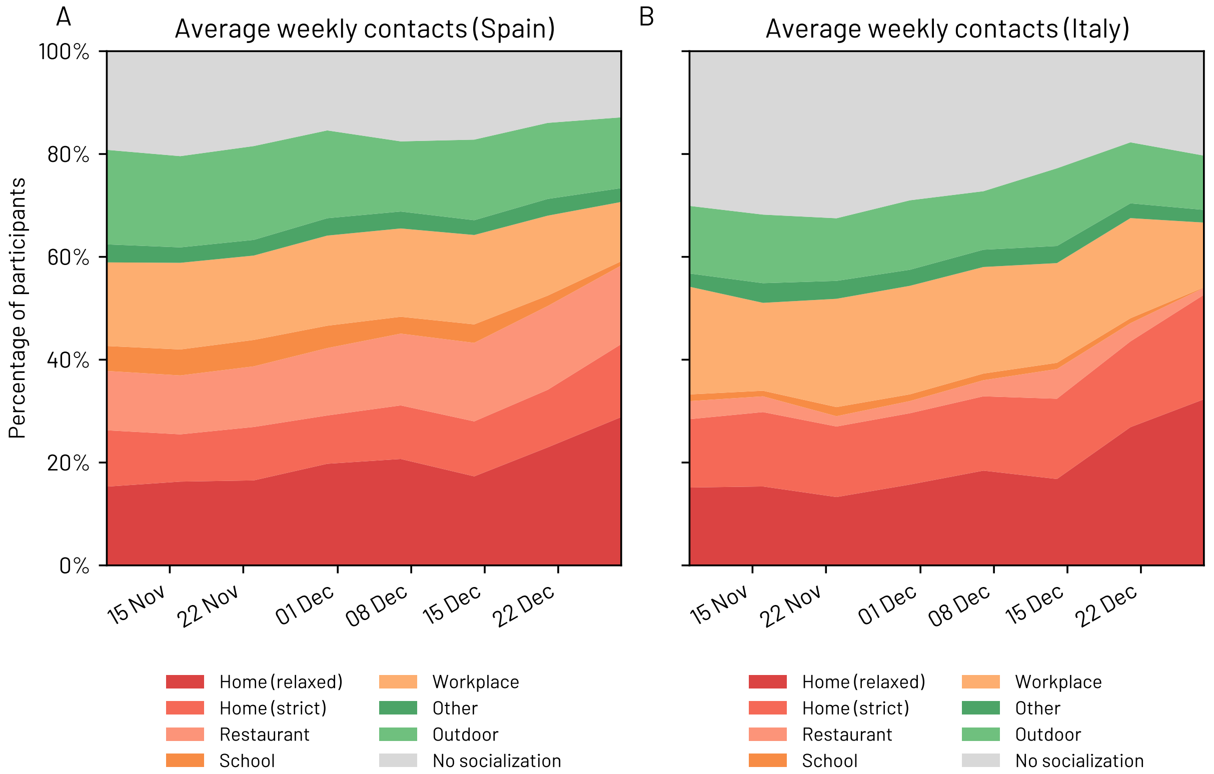


FIG. S16. Breakdown of the main environments of socialisation and their associated risk for Italy and Spain. The colours are chosen to ease the comparison between this Figure and the main paper Figure.

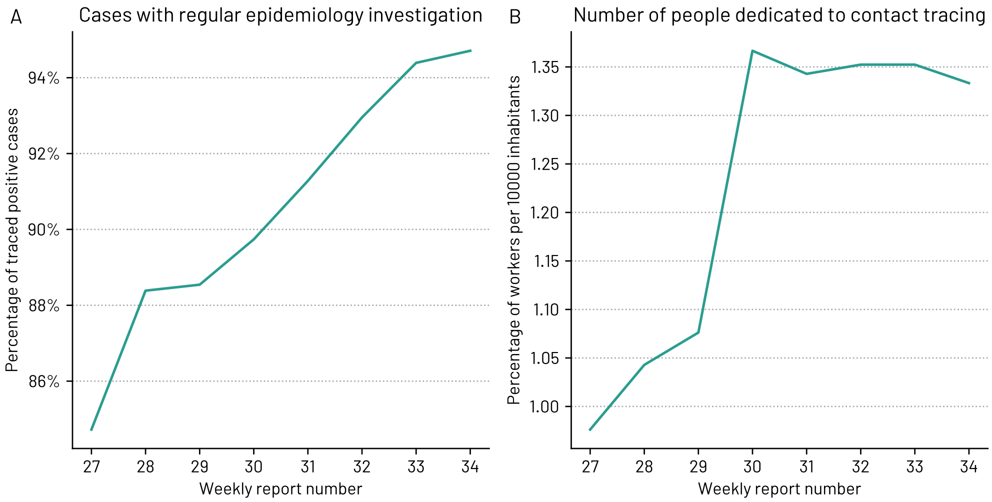


FIG. S17. Official data about contact tracing from the official reports of the Italy Health institute. Data shared by the Italian regions.

### S5. ADDITIONAL TABLES

| Country | Phase | Age range | | | | | | | |
|---|---|---|---|---|---|---|---|---|---|
| | | 18to20 | 21to29 | 30to39 | 40to49 | 50to59 | 60to69 | 70to79 | 80+ |
| Italy | Phase 1 | 452 | 2710 | 3236 | 3694 | 4408 | 4358 | 1559 | 155 |
| | Phase 2 | 1387 | 3846 | 4085 | 4024 | 4460 | 3679 | 1139 | 103 |
| Spain | Phase 1 | 1221 | 6808 | 11745 | 14258 | 15306 | 10783 | 3097 | 218 |
| | Phase 2 | 3502 | 5106 | 5283 | 5742 | 6064 | 4539 | 1247 | 90 |

TABLE S3. Number of answers per country, Phase and age group.

| Country | Work type | pre | post | change |
|---|---|---|---|---|
| Spain | Administrative | 14.16 | 19.52 | -27.46 |
| | Commercial | 4.23 | 4.02 | 5.34 |
| | Communications | 48.64 | 53.23 | -8.63 |
| | Construction | 2.37 | 4.83 | -50.92 |
| | Education | 6.70 | 27.85 | -75.95 |
| | Entertainment | 18.75 | 21.76 | -13.83 |
| | Essential services | 2.19 | 2.22 | -1.47 |
| | Finance | 27.22 | 34.76 | -21.68 |
| | Food production | 1.61 | 2.53 | -36.27 |
| | Health and social | 2.78 | 3.98 | -30.06 |
| | Hospitality | 1.23 | 1.43 | -13.67 |
| | Manufacturing | 3.15 | 5.44 | -42.02 |
| | Other services | 9.48 | 13.13 | -27.78 |
| | Public administration | 11.17 | 18.40 | -39.28 |
| | Science, Tech, Professional | 30.75 | 36.56 | -15.89 |
| | Transportation | 3.55 | 5.68 | -37.63 |
| Italy | Administrative | 16.11 | 19.54 | -17.55 |
| | Commercial | 5.97 | 1.82 | 228.44 |
| | Communications | 52.75 | 49.14 | 7.35 |
| | Construction | 9.86 | 3.12 | 215.57 |
| | Education | 28.77 | 24.56 | 17.18 |
| | Entertainment | 33.59 | 13.73 | 144.68 |
| | Essential services | 3.73 | 0.92 | 307.05 |
| | Finance | 37.78 | 31.01 | 21.85 |
| | Food production | 6.90 | 2.19 | 214.22 |
| | Health and social | 3.05 | 4.02 | -24.21 |
| | Hospitality | 2.31 | 0.71 | 226.95 |
| | Manufacturing | 7.21 | 8.55 | -15.69 |
| | Other services | 18.86 | 16.77 | 12.43 |
| | Public administration | 20.84 | 21.15 | -1.44 |
| | Science, Tech, Professional | 23.69 | 24.04 | -1.47 |
| | Transportation | 9.47 | 8.76 | 8.16 |

TABLE S4. Percentage of reported teleworking in Phase I (pre) and after the end of Phase I (post).

| Country | pre | post | change |
| --- | --- | --- | --- |
| Spain | 9.91 | 15.38 | -35.58 |
| Italy | 16.00 | 14.57 | 9.80 |

TABLE S5. Percentage of reported teleworking in Phase I (pre) and after the end of Phase I (post).

| Country | Work type | pre | post | change |
|---|---|---|---|---|
| Spain | Administrative | 4.48 | 4.50 | -0.51 |
| | Commercial | 4.88 | 4.79 | 1.79 |
| | Communications | 4.30 | 2.97 | 44.71 |
| | Construction | 6.64 | 5.21 | 27.61 |
| | Education | 2.79 | 5.60 | -50.11 |
| | Entertainment | 13.70 | 16.90 | -18.91 |
| | Essential services | 2.62 | 1.47 | 78.25 |
| | Finance | 2.80 | 1.41 | 99.20 |
| | Food production | 10.53 | 10.17 | 3.55 |
| | Health and social | 4.33 | 3.56 | 21.63 |
| | Hospitality | 20.18 | 14.21 | 42.07 |
| | Manufacturing | 4.88 | 5.47 | -10.66 |
| | Other services | 9.39 | 8.96 | 4.88 |
| | Public administration | 1.73 | 0.72 | 138.98 |
| | Science, Tech, Professional | 2.08 | 2.40 | -13.26 |
| | Transportation | 6.63 | 6.58 | 0.74 |
| Italy | Administrative | 3.27 | 1.94 | 68.85 |
| | Commercial | 5.20 | 3.35 | 55.07 |
| | Communications | 5.02 | 4.79 | 4.94 |
| | Construction | 3.17 | 5.84 | -45.72 |
| | Education | 3.11 | 5.68 | -45.31 |
| | Entertainment | 23.38 | 14.40 | 62.37 |
| | Essential services | 1.08 | 0.88 | 23.36 |
| | Finance | 1.04 | 1.25 | -16.66 |
| | Food production | 9.06 | 3.63 | 149.62 |
| | Health and social | 3.49 | 2.61 | 33.51 |
| | Hospitality | 25.20 | 9.13 | 175.91 |
| | Manufacturing | 2.80 | 1.54 | 81.67 |
| | Other services | 9.14 | 6.53 | 39.84 |
| | Public administration | 2.11 | 1.70 | 23.80 |
| | Science, Tech, Professional | 2.60 | 2.16 | 20.69 |
| | Transportation | 6.17 | 4.06 | 51.81 |

TABLE S6. Percentage of reported unemployed participants and respondents on unpaid leave in Phase I (pre) and after the end of Phase I (post).

| Country | Type | pre | post | change |
|---|---|---|---|---|
| Spain | Unpaid leave | 1.56 | 3.23 | -51.61 |
| | Lost job | 10.74 | 8.34 | 28.79 |
| Italy | Unpaid leave | 3.54 | 2.02 | 75.52 |
| | Lost job | 7.76 | 5.92 | 30.93 |

TABLE S7. Percentage of unemployed participants and respondents on unpaid leave in Phase I (pre) and after the end of Phase I (post).
